# A Randomized Placebo-Controlled Trial of Sarilumab in Hospitalized Patients with Covid-19

**DOI:** 10.1101/2021.05.13.21256973

**Authors:** Sumathi Sivapalasingam, David J. Lederer, Rafia Bhore, Negin Hajizadeh, Gerard Criner, Romana Hosain, Adnan Mahmood, Angeliki Giannelou, Selin Somersan-Karakaya, Meagan O’Brien, Anita Boyapati, Janie Parrino, Bret Musser, Emily Labriola-Tompkins, Divya Ramesh, Lisa A. Purcell, Daya Gulabani, Wendy Kampman, Alpana Waldron, Michelle Ng Gong, Suraj Saggar, Steven J. Sperber, Vidya Menon, David K. Stein, Magdalena E. Sobieszczyk, William Park, Judith A. Aberg, Samuel M. Brown, Jack A. Kosmicki, Julie E. Horowitz, Manuel A. Ferreira, Aris Baras, Bari Kowal, A. Thomas DiCioccio, Bolanle Akinlade, Michael C. Nivens, Ned Braunstein, Gary Herman, George D. Yancopoulos, David M. Weinreich, for the Sarilumab-COVID-19 Study Team

## Abstract

**BACKGROUND:** Sarilumab (anti-interleukin-6 receptor-α monoclonal antibody) may attenuate the inflammatory response in Covid-19.

**METHODS:** We performed an adaptive, phase 2/3, randomized, double-blind, placebo-controlled trial of intravenous sarilumab 200 mg or 400 mg in adults hospitalized with Covid-19. The phase 3 primary analysis population (cohort 1) was patients with critical Covid-19 receiving mechanical ventilation (MV) randomized to sarilumab 400 mg or placebo. The primary end point for phase 3 was the proportion of patients with ≥1-point improvement in clinical status from baseline to day 22.

**RESULTS:** Four-hundred fifty-seven (457) and 1365 patients were randomized and treated in phases 2 and 3, respectively. Among phase 3 critical patients receiving MV (n=289; 34.3% on corticosteroids), the proportion with ≥1-point improvement in clinical status (alive not receiving MV) at day 22 was 43.2% in sarilumab 400 mg and 35.5% in placebo (risk difference [RD] +7.5%; 95% confidence interval [CI], –7.4 to 21.3; P=0.3261), representing a relative risk improvement of 21.7%. Day 29 all-cause mortality was 36.4% in sarilumab 400 mg versus 41.9% in placebo (RD –5.5%; 95% CI, –20.2 to 8.7; relative risk reduction 13.3%). In post hoc analyses pooling phase 2 and 3 critical patients receiving MV, the hazard ratio (HR) for death in sarilumab 400 mg compared with placebo was 0.76 (95% CI, 0.51 to 1.13) overall, improving to 0.49 (95% CI, 0.25 to 0.94) in patients receiving corticosteroids at baseline.

**CONCLUSION:** In hospitalized patients with Covid-19 receiving MV, numerical benefits with sarilumab did not achieve statistical significance, but benefit may be greater in patients receiving corticosteroids. A larger study is required to confirm this observed numerical benefit.

(ClinicalTrials.gov number, NCT04315298)

## INTRODUCTION

Severe acute respiratory syndrome coronavirus 2 (SARS-CoV-2), a novel coronavirus first identified in December 2019,^1^ is the causative agent of coronavirus disease 2019 (Covid-19). While most patients with Covid-19 have mild disease, the leading cause of hospitalization and death is respiratory failure, including acute respiratory distress syndrome.^2,3^ A key driver of this deterioration may be a dysregulated inflammatory response^4^ based on the observation of elevated circulating levels of inflammatory mediators such as C-reactive protein (CRP) and interleukin-6 (IL-6).^5,6^ Early in the pandemic, small, uncontrolled studies reported that treating hospitalized patients with Covid-19 with two different IL-6 receptor (IL-6R) blocking antibodies, tocilizumab and sarilumab, resulted in potentially dramatic clinical improvement.^7-12^ More recently, two platform trials found that tocilizumab or sarilumab may improve clinical outcomes in patients with Covid-19, the majority of whom were also receiving corticosteroids.^13,14^

On March 13, 2020, Covid-19 was declared a national emergency in the United States.^15^ On March 18, 2020, we initiated a clinical trial to evaluate the efficacy and safety of intravenous (IV) sarilumab, an anti–IL-6R monoclonal antibody, for the treatment of hospitalized patients with Covid-19 in the United States (ClinicalTrials.gov number, NCT04315298). Here we report the final results of this trial.

## METHODS

### TRIAL DESIGN, PATIENTS, AND TREATMENTS

We conducted an adaptive, phase 2/3, randomized, double-blind, placebo-controlled, multicenter trial (65 sites in total, with 62 enrolling patients; see full list of sites in the **Supplementary Appendix**). Since the trial was being conducted during a pandemic with a novel coronavirus, the protocol allowed adaptations including closing treatment arms or randomization strata, modification of the provisional phase 3 end points, and sample size re-estimation before phase 3 database lock and study readout.

Eligible patients were ≥18 years of age and hospitalized with laboratory-confirmed SARS-CoV-2 infection requiring supplemental oxygen and/or assisted ventilation. All patients received local standard of care (SOC), including corticosteroids and open-label use of putative treatments for Covid-19 (see **Supplementary Appendix**).

In phase 2, patients were randomized in a 2:2:1 ratio to IV sarilumab 400 mg, sarilumab 200 mg, or placebo (instructions on dose preparation provided in the **Supplementary Appendix**). Randomization was stratified by corticosteroid use and disease severity (severe, critical, multi-system organ dysfunction [MSOD], and immunocompromised state). Further details provided in the **Supplementary Appendix**.

### TRIAL ADAPTATIONS

A summary of protocol amendments and study adaptations are found **Supplementary Appendix**. The phase 2 and phase 3 parts of the study used the same entry and stratification criteria (**Fig. S1**) until a pre-specified phase 2 interim analysis of 457 patients indicated potential benefit of sarilumab 400 mg in patients in the critical stratum (high-flow supplemental oxygen and/or mechanical ventilation) and potential harm of sarilumab 400 mg in patients in the severe (low-flow supplemental oxygen) and MSOD strata (see **Supplementary Appendix** for complete definitions of disease severity strata). Subsequently, the Independent Data Monitoring Committee recommended discontinuation of enrollment into the severe and MSOD strata and elimination of the 200-mg dose. Thereafter, the phase 3 protocol was amended to restrict enrollment to critical patients receiving mechanical ventilation with further randomization (2:1) to sarilumab 400 mg and placebo (cohort 1; **Fig. S2**) and to add a new cohort of critical patients receiving mechanical ventilation randomized to sarilumab 800 mg or placebo (cohort 2) and a new cohort of critical patients not receiving mechanical ventilation, but requiring high-flow oxygen or non-invasive ventilation, randomized to sarilumab 800 mg or placebo (cohort 3). In addition, the following adaptations were implemented prior to database lock: the phase 3 primary analysis population was changed to patients randomized to the critical stratum receiving MV (without extracorporeal membrane oxygenation [ECMO]) who were randomized to the sarilumab 400-mg group or placebo, the primary end point was changed from the time to ≥2-point improvement in clinical status using a 7-point ordinal scale to the proportion with ≥1-point improvement from baseline to day 22, and the sample size was re-calculated.

Results from the phase 3 immunocompromised stratum (n=35 patients randomized and treated) are not shown; results for cohorts 2 and 3 (sarilumab 800 mg) are reported in the **Supplementary Appendix**.

### END POINTS

The pre-specified phase 2 primary end point was the percent change from baseline in CRP level at day 4. Following the interim analysis from the phase 2 part of the study, the pre-specified primary end point for phase 3 cohort 1 was the proportion of critical patients receiving MV at baseline with ≥1-point improvement in clinical status on a 7-point ordinal scale from baseline to day 22 (Clinical Status Scale in the **Supplementary Appendix**).^16^ In this population, a 1-point improvement indicates that the patient was alive and no longer receiving MV. Other pre-specified end points included the proportion of patients who died by day 60 and the proportion of patients who recovered (discharged or alive without supplemental oxygen use) by day 22. All outcomes were assessed by the site investigators, who were blinded to treatment assignment and serum IL-6 levels conducted at a central laboratory. Safety end points are described in the **Supplementary Appendix**.

### STUDY OVERSIGHT

Details are provided in the **Supplementary Appendix**.

### STATISTICAL ANALYSIS

The primary analysis population for phase 2 included all randomized patients who received study drug and had high baseline IL-6 levels (above the upper limit of normal) in all disease strata. The primary analysis population for phase 3 included all patients with critical Covid-19 receiving MV without ECMO at baseline who were randomized to the sarilumab 400-mg group or placebo group. Patients in phase 2 were not included in the phase 3 portion of the study.

Hypothesis tests of superiority of sarilumab 400 mg versus placebo were performed with the stratified Cochran–Mantel–Haenszel (CMH) test for two proportions. The stratification factor was the use of corticosteroids at baseline. P values and strata-adjusted confidence intervals (CIs) from the CMH test were reported with overall type 1 error controlled at 0.05 (two-sided). The primary and key secondary end points in the phase 3 study were tested sequentially in a hierarchical manner, while preserving the overall significance level at 0.05 (two-sided). Additional statistical details are provided in the **Supplementary Appendix** and the statistical analysis plan.

Post hoc analyses were conducted in the pooled data from phase 2 and phase 3 (cohort 1) to descriptively analyze the end points of the proportion of patients with ≥1-point improvement in clinical status to day 22, proportion alive and not receiving MV, and time to death. All post hoc analyses were unstratified.

## RESULTS

From the period of March 18, 2020, to July 2, 2020, 457 patients were randomized and treated in phase 2 (placebo, n=90; sarilumab 200 mg, n=187; sarilumab 400 mg, n=180), and 1365 patients were randomized and treated in phase 3 cohort 1 (placebo, n=294; sarilumab 200 mg, n=489; sarilumab 400 mg, n=582; **Fig. S2**). A summary of phase 2 and phase 3 analysis populations by disease severity strata is presented in **Table S1**. Results of the phase 2 portion of the study are presented in the **Supplementary Appendix**.

### BASELINE DEMOGRAPHICS AND CHARACTERISTICS

In phase 3, 750 (54.9%) patients were randomized to the critical stratum (298 [21.8%] receiving MV at baseline, 450 [33.0%] were not receiving MV, 2 [0.3%] receiving ECMO); 347 (25.4%) were randomized to the severe stratum, and 233 (17.1%) to the MSOD stratum. The median age of patients in the critical stratum was 61 years, 68% were male, and the median duration of illness was 9.0 days. The median serum CRP, serum IL-6 concentration, and viral load in nasopharyngeal swabs were 168.80 mg/l, 110.91 pg/ml, and 3.95 log_10_ copies/ml, respectively. At randomization, 34.3% of critical patients were receiving systemic corticosteroids, and this was balanced across treatment groups (**Table 1**).

**Table 1.**
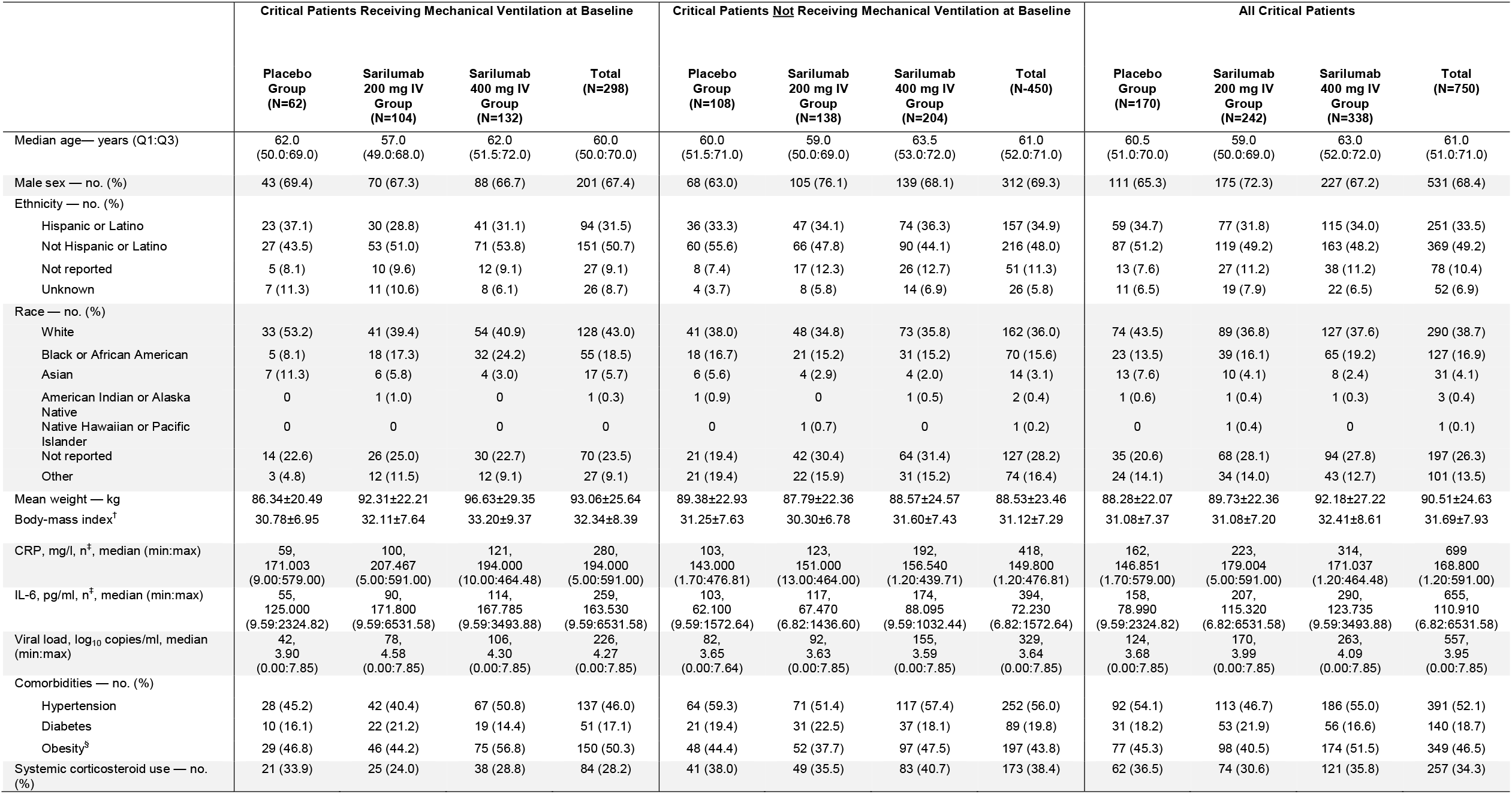
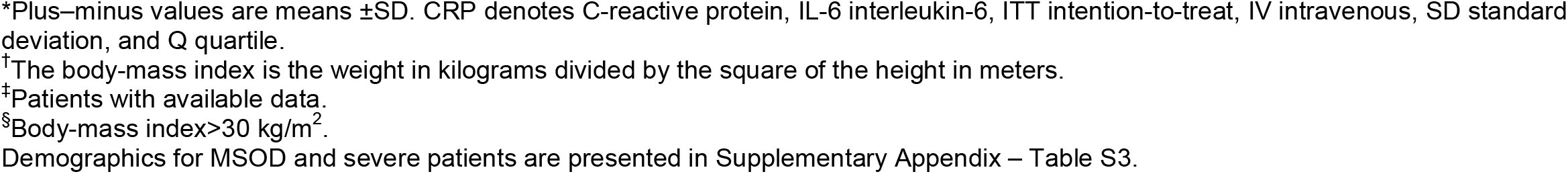
Phase 3 (Cohort 1) – Critical Stratum: Demographics and Baseline Characteristics (ITT Population).*

|  | Critical Patients Receiving Mechanical Ventilation at Baseline |  |  |  | Critical Patients <u>Not</u> Receiving Mechanical Ventilation at Baseline |  |  |  | All Critical Patients |  |  |  |
| --- | --- | --- | --- | --- | --- | --- | --- | --- | --- | --- | --- | --- |
|  | Placebo Group (N=62) | Sarilumab 200 mg IV Group (N=104) | Sarilumab 400 mg IV Group (N=132) | Total (N=298) | Placebo Group (N=108) | Sarilumab 200 mg IV Group (N=138) | Sarilumab 400 mg IV Group (N=204) | Total (N=450) | Placebo Group (N=170) | Sarilumab 200 mg IV Group (N=242) | Sarilumab 400 mg IV Group (N=338) | Total (N=750) |
| Median age— years (Q1:Q3) | 62.0<br>(50.0:69.0) | 57.0<br>(49.0:68.0) | 62.0<br>(51.5:72.0) | 60.0<br>(50.0:70.0) | 60.0<br>(51.5:71.0) | 59.0<br>(50.0:69.0) | 63.5<br>(53.0:72.0) | 61.0<br>(52.0:71.0) | 60.5<br>(51.0:70.0) | 59.0<br>(50.0:69.0) | 63.0<br>(52.0:72.0) | 61.0<br>(51.0:71.0) |
| Male sex — no. (%) | 43 (69.4) | 70 (67.3) | 88 (66.7) | 201 (67.4) | 68 (63.0) | 105 (76.1) | 139 (68.1) | 312 (69.3) | 111 (65.3) | 175 (72.3) | 227 (67.2) | 531 (68.4) |
| Ethnicity — no. (%) |  |  |  |  |  |  |  |  |  |  |  |  |
| Hispanic or Latino | 23 (37.1) | 30 (28.8) | 41 (31.1) | 94 (31.5) | 36 (33.3) | 47 (34.1) | 74 (36.3) | 157 (34.9) | 59 (34.7) | 77 (31.8) | 115 (34.0) | 251 (33.5) |
| Not Hispanic or Latino | 27 (43.5) | 53 (51.0) | 71 (53.8) | 151 (50.7) | 60 (55.6) | 66 (47.8) | 90 (44.1) | 216 (48.0) | 87 (51.2) | 119 (49.2) | 163 (48.2) | 369 (49.2) |
| Not reported | 5 (8.1) | 10 (9.6) | 12 (9.1) | 27 (9.1) | 8 (7.4) | 17 (12.3) | 26 (12.7) | 51 (11.3) | 13 (7.6) | 27 (11.2) | 38 (11.2) | 78 (10.4) |
| Unknown | 7 (11.3) | 11 (10.6) | 8 (6.1) | 26 (8.7) | 4 (3.7) | 8 (5.8) | 14 (6.9) | 26 (5.8) | 11 (6.5) | 19 (7.9) | 22 (6.5) | 52 (6.9) |
| Race — no. (%) |  |  |  |  |  |  |  |  |  |  |  |  |
| White | 33 (53.2) | 41 (39.4) | 54 (40.9) | 128 (43.0) | 41 (38.0) | 48 (34.8) | 73 (35.8) | 162 (36.0) | 74 (43.5) | 89 (36.8) | 127 (37.6) | 290 (38.7) |
| Black or African American | 5 (8.1) | 18 (17.3) | 32 (24.2) | 55 (18.5) | 18 (16.7) | 21 (15.2) | 31 (15.2) | 70 (15.6) | 23 (13.5) | 39 (16.1) | 65 (19.2) | 127 (16.9) |
| Asian | 7 (11.3) | 6 (5.8) | 4 (3.0) | 17 (5.7) | 6 (5.6) | 4 (2.9) | 4 (2.0) | 14 (3.1) | 13 (7.6) | 10 (4.1) | 8 (2.4) | 31 (4.1) |
| American Indian or Alaska Native | 0 | 1 (1.0) | 0 | 1 (0.3) | 1 (0.9) | 0 | 1 (0.5) | 2 (0.4) | 1 (0.6) | 1 (0.4) | 1 (0.3) | 3 (0.4) |
| Native Hawaiian or Pacific Islander | 0 | 0 | 0 | 0 | 0 | 1 (0.7) | 0 | 1 (0.2) | 0 | 1 (0.4) | 0 | 1 (0.1) |
| Not reported | 14 (22.6) | 26 (25.0) | 30 (22.7) | 70 (23.5) | 21 (19.4) | 42 (30.4) | 64 (31.4) | 127 (28.2) | 35 (20.6) | 68 (28.1) | 94 (27.8) | 197 (26.3) |
| Other | 3 (4.8) | 12 (11.5) | 12 (9.1) | 27 (9.1) | 21 (19.4) | 22 (15.9) | 31 (15.2) | 74 (16.4) | 24 (14.1) | 34 (14.0) | 43 (12.7) | 101 (13.5) |
| Mean weight — kg | 86.34±20.49 | 92.31±22.21 | 96.63±29.35 | 93.06±25.64 | 89.38±22.93 | 87.79±22.36 | 88.57±24.57 | 88.53±23.46 | 88.28±22.07 | 89.73±22.36 | 92.18±27.22 | 90.51±24.63 |
| Body-mass index<sup>†</sup> | 30.78±6.95 | 32.11±7.64 | 33.20±9.37 | 32.34±8.39 | 31.25±7.63 | 30.30±6.78 | 31.60±7.43 | 31.12±7.29 | 31.08±7.37 | 31.08±7.20 | 32.41±8.61 | 31.69±7.93 |
| CRP, mg/l, n<sup>‡</sup>, median (min:max) | 59,<br>171.003<br>(9.00:579.00) | 100,<br>207.467<br>(5.00:591.00) | 121,<br>194.000<br>(10.00:464.48) | 280,<br>194.000<br>(5.00:591.00) | 103,<br>143.000<br>(1.70:476.81) | 123,<br>151.000<br>(13.00:464.00) | 192,<br>156.540<br>(1.20:439.71) | 418,<br>149.800<br>(1.20:476.81) | 162,<br>146.851<br>(1.70:579.00) | 223,<br>179.004<br>(5.00:591.00) | 314,<br>171.037<br>(1.20:464.48) | 699,<br>168.800<br>(1.20:591.00) |
| IL-6, pg/ml, n<sup>‡</sup>, median (min:max) | 55,<br>125.000<br>(9.59:2324.82) | 90,<br>171.800<br>(9.59:6531.58) | 114,<br>167.785<br>(9.59:3493.88) | 259,<br>163.530<br>(9.59:6531.58) | 103,<br>62.100<br>(9.59:1572.64) | 117,<br>67.470<br>(6.82:1436.60) | 174,<br>88.095<br>(9.59:1032.44) | 394,<br>72.230<br>(6.82:1572.64) | 158,<br>78.990<br>(9.59:2324.82) | 207,<br>115.320<br>(6.82:6531.58) | 290,<br>123.735<br>(9.59:3493.88) | 655,<br>110.910<br>(6.82:6531.58) |
| Viral load, log<sub>10</sub> copies/ml, median (min:max) | 42,<br>3.90<br>(0.00:7.85) | 78,<br>4.58<br>(0.00:7.85) | 106,<br>4.30<br>(0.00:7.85) | 226,<br>4.27<br>(0.00:7.85) | 82,<br>3.65<br>(0.00:7.64) | 92,<br>3.63<br>(0.00:7.85) | 155,<br>3.59<br>(0.00:7.85) | 329,<br>3.64<br>(0.00:7.85) | 124,<br>3.68<br>(0.00:7.85) | 170,<br>3.99<br>(0.00:7.85) | 263,<br>4.09<br>(0.00:7.85) | 557,<br>3.95<br>(0.00:7.85) |
| Comorbidities — no. (%) |  |  |  |  |  |  |  |  |  |  |  |  |
| Hypertension | 28 (45.2) | 42 (40.4) | 67 (50.8) | 137 (46.0) | 64 (59.3) | 71 (51.4) | 117 (57.4) | 252 (56.0) | 92 (54.1) | 113 (46.7) | 186 (55.0) | 391 (52.1) |
| Diabetes | 10 (16.1) | 22 (21.2) | 19 (14.4) | 51 (17.1) | 21 (19.4) | 31 (22.5) | 37 (18.1) | 89 (19.8) | 31 (18.2) | 53 (21.9) | 56 (16.6) | 140 (18.7) |
| Obesity<sup>§</sup> | 29 (46.8) | 46 (44.2) | 75 (56.8) | 150 (50.3) | 48 (44.4) | 52 (37.7) | 97 (47.5) | 197 (43.8) | 77 (45.3) | 98 (40.5) | 174 (51.5) | 349 (46.5) |
| Systemic corticosteroid use — no. (%) | 21 (33.9) | 25 (24.0) | 38 (28.8) | 84 (28.2) | 41 (38.0) | 49 (35.5) | 83 (40.7) | 173 (38.4) | 62 (36.5) | 74 (30.6) | 121 (35.8) | 257 (34.3) |

\*Plus–minus values are means ± SD. CRP denotes C-reactive protein, IL-6 interleukin-6, ITT intention-to-treat, IV intravenous, SD standard deviation, and Q quartile.
<sup>†</sup>The body-mass index is the weight in kilograms divided by the square of the height in meters.
<sup>‡</sup>Patients with available data.
<sup>§</sup>Body-mass index $>30$ kg/m<sup>2</sup>.
Demographics for MSOD and severe patients are presented in Supplementary Appendix – Table S3.

Demographics and baseline characteristics for phase 3 severe and MSOD strata and for the pooled phase 2/3 dataset are presented in **Tables S2** and **S3**, respectively.

### PHASE 3 EFFICACY OUTCOMES

#### Primary Outcome

In the pre-specified primary analysis among critical patients receiving MV, 43.2% of patients in the 400-mg group and 35.5% patients in the placebo group had ≥1-point improvement in clinical status (alive not receiving MV) at day 22 (risk difference [RD] +7.5%; 95% CI, –7.4 to 21.3; P=0.3261; **Table 2**; **Fig. 1A**), representing a relative risk improvement of 21.7%.

**Table 2.** Phase 3 (Cohort 1): Primary Efficacy End Point.*

| Proportion of patients with ≥1-point improvement in clinical status from baseline to day 22<br>(critical ITT patients receiving mechanical ventilation at baseline) |  |  |  |
| --- | --- | --- | --- |
|  | Placebo Group<br>(N=62) | Sarilumab 200 mg IV Group<br>(N=104) | Sarilumab 400 mg IV Group<br>(N=132) |
| no. – (%) | 22 (35.5) | 45 (43.3) | 57 (43.2) |
| 95% CI | 23.6–47.4 | 33.7–52.8 | 34.7–51.6 |
| Risk difference vs. placebo (%) | - | 7.1% | 7.5% |
| 95% CI | - | -8.4%–21.7% | -7.4%–21.3% |
| P value | - | 0.3707 | 0.3261 |

\*CI denotes confidence interval, ITT intention-to-treat, IV intravenous.

**Figure 1.**
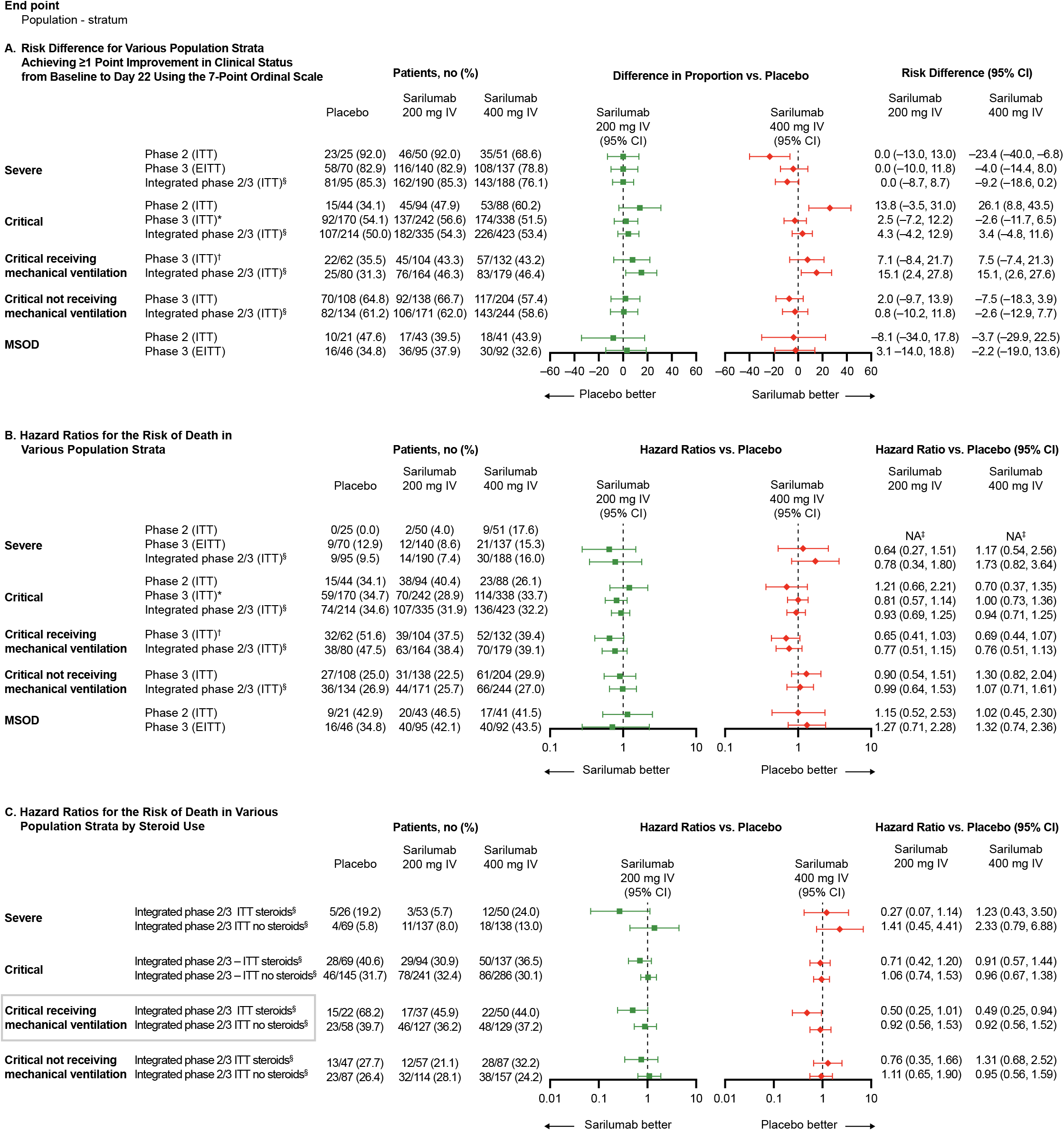
A) Risk difference for various population strata achieving ≥1 point improvement in clinical status from baseline to Day 22 using the 7-point ordinal scale; B) hazard ratios for the risk of death in various population strata; C) hazard ratios for the risk of death in various population strata by steroid use. *Key secondary endpoint – phase 3 (cohort 1) †Primary endpoint – phase 3 (cohort 1) ‡Hazard ratio could not be calculated, as the number of events was too small. §Unstratified analysis. CI, confidence interval; EITT, exploratory intention-to-treat; ITT, intention-to-treat; IV, intravenous; MSOD, multi-systemic organ dysfunction.

#### Secondary Outcomes

Among critical patients receiving MV at baseline, mortality by day 29 was 36.4% in the 400-mg group and 41.9% in the placebo group (RD –5.5%; 95% CI, –20.2 to 8.7; relative risk reduction 13.3%; key secondary end point) and mortality by day 60 was 39.4% in the sarilumab 400-mg group and 51.6% in the placebo group (RD –11.9%; 95% CI, –26.4 to 2.9; relative risk reduction 23.7%; **Table S4**). Time to death in critical patients receiving MV at baseline is shown in **Figure 2**. Recovery by day 22 occurred in 31.8% of critical patients receiving MV at baseline in the 400-mg group and 25.8% in the placebo group (RD +5.7%; 95% CI, –8.4 to 18.2; relative risk reduction 23.3%; key secondary end point; **Table S4**).

**Figure 2.**
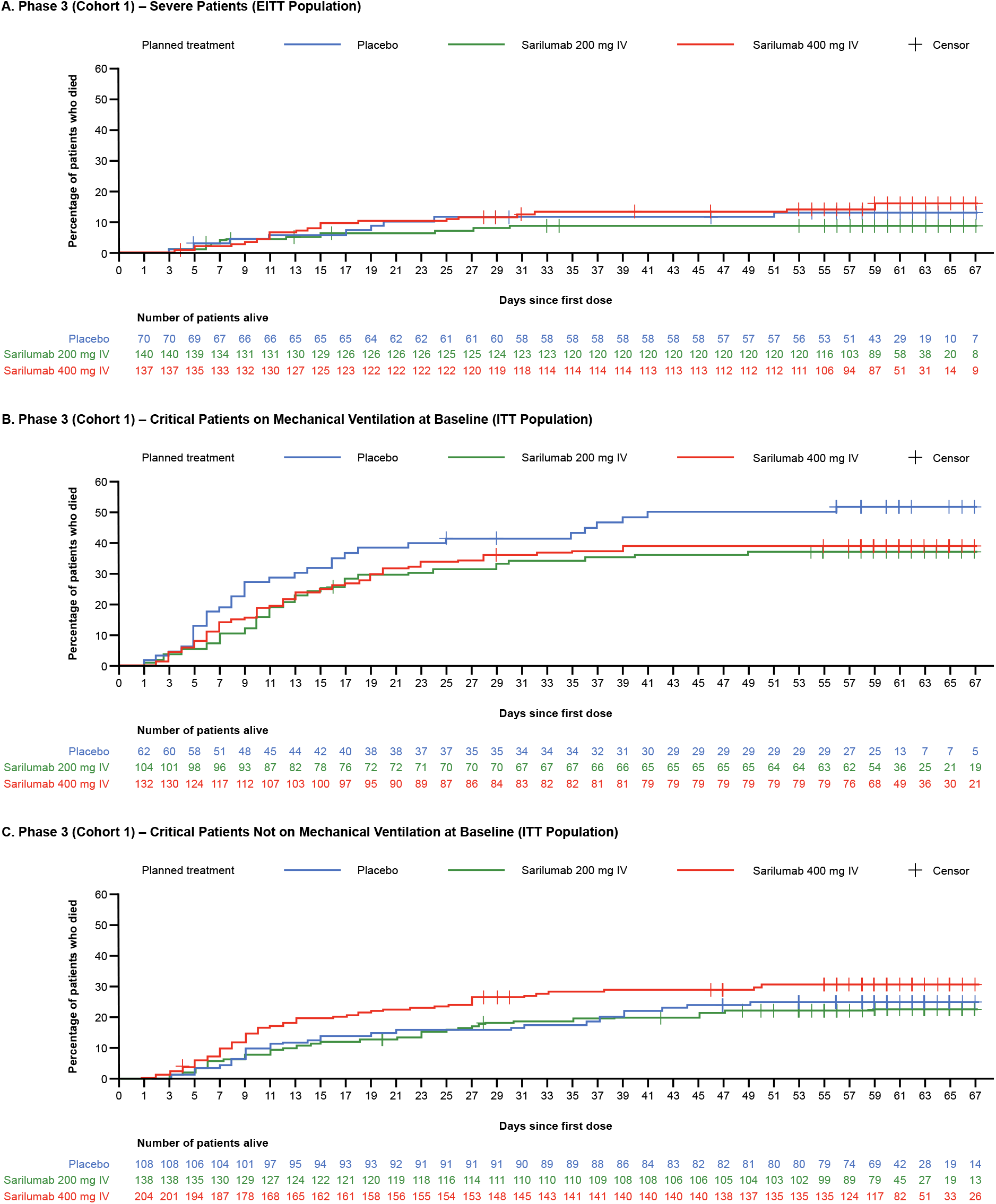
Kaplan Meier curve of time to death in the phase 3 (cohort 1) for A) severe patients (EITT Population); B) critical patients on mechanical ventilation at baseline (ITT Population); C) critical patients not on mechanical ventilation at baseline (ITT Population).

Among critical patients not receiving MV at baseline, 57.4% of patients in the 400-mg group and 64.8% of patients in the placebo group had ≥1-point improvement in clinical status at day 22 (RD –7.5%; 95% CI, –18.3 to 3.9; **Table S4**). Mortality by day 29 was 26.5% in the 400-mg group and 15.7% in the placebo group (RD +10.7%; 95% CI, 0.9 to 19.3; **Table S4**), and mortality by day 60 was 29.9% in the 400-mg group and 25% in the placebo group (RD +4.8%; 95% CI, 5.8 to 14.6; **Table S4**). Time to death in critical patients not receiving MV at baseline is shown in **Figure 2**. Recovery by day 22 occurred in 54.4% of patients in the 400-mg group and 63.9% in the placebo group (RD –9.5%; 95% CI, –20.4 to 2.0; **Table S4**).

#### Prespecified exploratory analyses

There was no evidence of efficacy of sarilumab compared with placebo in severe or MSOD patients (**Table S5**; **Fig. 1**; **Fig. 2A**).

### POST HOC ANALYSES

In post hoc analyses of pooled phase 2 and phase 3 critical patients receiving MV, 46.4% of patients receiving 400 mg and 31.3% of patients receiving placebo had ≥1-point improvement in clinical status at day 22 (RD 15.1%; 95% CI, 2.6 to 27.6; **Fig. 1A**). Similar findings were observed for mortality (**Fig. 1B**). No benefit in clinical status was seen in critical patients not receiving MV at baseline (RD –2.6%; 95% CI, –12.9 to 7.7; **Fig. 1A**).

Among critical patients receiving MV and corticosteroids at baseline, 44.0% (22 of 50) of patients died in the 400 mg-group compared with 68.2% (15 of 22) in the placebo group (HR, 0.49; 95% CI, 0.25 to 0.94; **Fig. 1C**). Among critical patients receiving MV without corticosteroids at baseline, 37.2% (48 of 129) died in the 400 mg-group compared with 39.7% (23 of 58) in the placebo group (HR, 0.92, 95% CI, 0.56 to 1.52; **Fig. 1C**).

### SAFETY

In the phase 3 portion of the study in critical patients, the safety profiles of sarilumab 200 mg and 400 mg were comparable and were similar to placebo (**Table S6**). Adverse events were consistent with advanced Covid-19 and its complications. Serious adverse events (SAEs) of cardiac arrest, Covid-19, respiratory failure, acute respiratory failure, septic shock, and acute kidney injury were reported in a similar proportion of patients in the sarilumab dose groups in comparison with placebo group, suggesting that these events were related to Covid-19 (**Table 3**).

**Table 3.** Phase 3 (Cohort 1) – Critical Stratum: Treatment-Emergent Serious Adverse Events and Adverse Events of Special Interest Occurring in ≥5% of Patients in Any Group (Safety Population).*

| System Organ Class<br>Preferred Term | Critical Patients Receiving Mechanical<br>Ventilation at Baseline |  |  | Critical Patients <u>Not</u> Receiving Mechanical<br>Ventilation at Baseline |  |  | All Critical Patients |  |  |
| --- | --- | --- | --- | --- | --- | --- | --- | --- | --- |
|  | Placebo<br>(N=62) | Sarilumab<br>200 mg IV<br>Group<br>(N=104) | Sarilumab<br>400 mg IV<br>Group<br>(N=132) | Placebo<br>(N=108) | Sarilumab<br>200 mg IV<br>Group<br>(N=138) | Sarilumab<br>400 mg IV<br>Group<br>(N=204) | Placebo<br>(N=170) | Sarilumab<br>200 mg IV<br>Group<br>(N=242) | Sarilumab<br>400 mg IV<br>Group<br>(N=338) |
| **SAEs** |  |  |  |  |  |  |  |  |  |
| SAEs — no. | 103 | 143 | 174 | 96 | 114 | 183 | 199 | 257 | 359 |
| Patients with ≥1 SAE — no. (%) | 44 (71.0) | 65 (62.5) | 88 (66.7) | 45 (41.7) | 54 (39.1) | 93 (45.6) | 89 (52.4) | 119 (49.2) | 183 (54.1) |
| Infections and infestations — no. (%) |  |  |  |  |  |  |  |  |  |
| Covid-19 | 6 (9.7) | 9 (8.7) | 9 (6.8) | 9 (8.3) | 10 (7.2) | 14 (6.9) | 15 (8.8) | 19 (7.9) | 23 (6.8) |
| Septic shock | 6 (9.7) | 5 (4.8) | 6 (4.5) | 3 (2.8) | 1 (0.7) | 6 (2.9) | 9 (5.3) | 6 (2.5) | 12 (3.6) |
| Respiratory, thoracic, and mediastinal<br>disorders — no. (%) |  |  |  |  |  |  |  |  |  |
| Respiratory failure | 5 (8.1) | 9 (8.7) | 7 (5.3) | 10 (9.3) | 14 (10.1) | 14 (6.9) | 15 (8.8) | 23 (9.5) | 22 (6.5) |
| Acute respiratory failure | 3 (4.8) | 4 (3.8) | 5 (3.8) | 7 (6.5) | 2 (1.4) | 12 (5.9) | 10 (5.9) | 6 (2.5) | 17 (5.0) |
| Pneumothorax | 2 (3.2) | 6 (5.8) | 1 (0.8) | 3 (2.8) | 2 (1.4) | 8 (3.9) | 5 (2.9) | 8 (3.3) | 9 (2.7) |
| Cardiac disorders — no. (%) |  |  |  |  |  |  |  |  |  |
| Cardiac arrest | 5 (8.1) | 6 (5.8) | 11 (8.3) | 2 (1.9) | 1 (0.7) | 14 (6.9) | 7 (4.1) | 7 (2.9) | 25 (7.4) |
| Renal and urinary disorders — no. (%) |  |  |  |  |  |  |  |  |  |
| Acute kidney injury | 6 (9.7) | 9 (8.7) | 3 (2.3) | 4 (3.7) | 3 (2.2) | 7 (3.4) | 10 (5.9) | 12 (5.0) | 10 (3.0) |
| General disorders and administration<br>site conditions — no. (%) |  |  |  |  |  |  |  |  |  |
| Multiple organ dysfunction syndrome | 3 (4.8) | 1 (1.0) | 8 (6.1) | 1 (0.9) | 3 (2.2) | 2 (1.0) | 4 (2.4) | 4 (1.7) | 10 (3.0) |
| **AESIs** |  |  |  |  |  |  |  |  |  |
| AESIs — no. | 61 | 102 | 128 | 49 | 89 | 124 | 110 | 191 | 255 |
| Patients with ≥1 AESI — no. (%) | 31 (50.0) | 56 (53.8) | 71 (53.8) | 30 (27.8) | 51 (37.0) | 77 (37.7) | 61 (35.9) | 107 (44.2) | 150 (44.4) |
| Investigations — no. (%) |  |  |  |  |  |  |  |  |  |
| Alanine aminotransferase increased | 8 (12.9) | 23 (22.1) | 16 (12.1) | 13 (12.0) | 23 (16.7) | 37 (18.1) | 21 (12.4) | 46 (19.0) | 55 (16.3) |
| Aspartate aminotransferase<br>increased | 8 (12.9) | 21 (20.2) | 18 (13.6) | 12 (11.1) | 19 (13.8) | 19 (9.3) | 20 (11.8) | 40 (16.5) | 38 (11.2) |
| Transaminases increased | 3 (4.8) | 4 (3.8) | 12 (9.1) | 3 (2.8) | 3 (2.2) | 16 (7.8) | 6 (3.5) | 7 (2.9) | 28 (8.3) |
| Liver function test increased | 2 (3.2) | 2 (1.9) | 8 (6.1) | 0 (0.0) | 1 (0.7) | 0 (0.0) | 3 (1.8) | 2 (0.8) | 10 (3.0) |
| Infections and infestations — no. (%) |  |  |  |  |  |  |  |  |  |
| Pneumonia | 4 (6.5) | 3 (2.9) | 8 (6.1) | 2 (1.9) | 1 (0.7) | 3 (1.5) | 6 (3.5) | 4 (1.7) | 11 (3.3) |
| Staphylococcal infection | 4 (6.5) | 2 (1.9) | 5 (3.8) | 2 (1.9) | 2 (1.4) | 3 (1.5) | 6 (3.5) | 4 (1.7) | 8 (2.4) |

\*AESI denotes adverse event of special interest, IV intravenous, SAE serious adverse event.

Adverse events of special interest (AESIs) reported in more than 5% of the patients in any treatment group were elevations of liver function tests, reported numerically more often in patients receiving sarilumab than placebo (**Table 3**). None of the liver transaminase elevation cases met Hy’s law criteria.

In the phase 3 portion of the study, patients with severe disease or MSOD exhibited a similar safety profile to critical patients, and no new safety findings were observed (**Table S7**). Numerically, more patients reported elevation of liver function tests and SAEs in the sarilumab 200-mg and 400-mg dose groups in comparison with placebo (**Table S8**), which is expected, based on the known safety profile of sarilumab.^17^

### HUMAN GENETIC STUDIES

To independently explore the association between IL-6 signaling and Covid-19 outcomes, we focused on a common partial loss of function allele in the *IL6R* gene (rs2228145:C, 39% allele frequency in Europeans) that is associated with 7% lower CRP levels,^18^ reduced surface IL-6R levels,^19^ 50% higher IL-6R shedding,^20^ and 34% higher serum protein levels of soluble IL-6R (sIL-6R) (see Human Genetic Studies Methods in the **Supplementary Appendix**). This allele reduces risk of rheumatoid arthritis by 7%^21^ and coronary artery disease by 5%,^22^ while it increases the risk of asthma by 9%^23^ and atopic dermatitis by 15%.^24^

Using analysis from the COVID-19 Host Genetics Initiative,^25^ we first assessed the association between the partial loss of function variant (rs2228145:C) and risk of hospitalization, comparing individuals with inpatient hospitalization and polymerase chain reaction (PCR)–confirmed SARS-CoV-2 infection (n=12,888) against population-level controls with either a negative PCR test result or no PCR test (n=1,295,966). We observed a nominal association between the partial loss of function allele (rs2228145:C) and reduced odds of hospitalization (pan-ancestry meta-analysis per-allele odds ratio, 0.96; 95% CI, 0.93 to 0.99; P=0.007; **Table S9**).

## DISCUSSION

In this phase 2/3 clinical trial conducted early during the Covid-19 pandemic, sarilumab did not lead to significant improvement in clinical status or mortality in hospitalized patients with Covid-19, the majority of whom were not receiving corticosteroids as SOC. However, in phase 3 cohort 1, a numerically lower proportion of critical patients receiving mechanical ventilation in the sarilumab 400 mg had died by day 29 compared with placebo (36.4% vs 41.9%, respectively), a smaller effect than that observed with dexamethasone in the RECOVERY trial.^26^ There was also no benefit of sarilumab seen in patients receiving either low-or high-flow supplemental oxygen or non-invasive ventilation. Because only a minority of patients in our trial (∼30%) were receiving concomitant corticosteroids, these overall outcomes may not be generalizable to the hospitalized population receiving sarilumab in addition to corticosteroids as part of SOC.

In our post hoc analyses where we pooled phase 2 and 3 datasets, critical patients receiving MV at baseline had a mortality rate of 68% with steroids alone (placebo group) compared with 44% with steroids and sarilumab 400 mg. An additive benefit of anti–IL-6R therapy to corticosteroids in patients with Covid-19 has previously been reported in two platform trials. The REMAP-CAP trial, in which >90% of patients were receiving corticosteroids as part of SOC, reported a mortality benefit of tocilizumab and sarilumab compared with SOC alone in hospitalized patients requiring organ support.^13^ The RECOVERY trial in hospitalized patients with Covid-19 with hypoxemia and CRP >75 mg/l, in which 82% of patients were receiving corticosteroids as SOC, demonstrated that tocilizumab led to a statistically significant reduction in mortality (29% in the tocilizumab group versus 33% in SOC only). Tocilizumab treatment of patients not receiving mechanical ventilation reduced the risk of progressing to mechanical ventilation or death (33% vs. 38% SOC). Similar to the REMAP-CAP trial, the majority (82%) of patients were receiving corticosteroids at baseline, and the mortality benefit of tocilizumab was not seen in patients not receiving corticosteroids.^14^

Discussion of the results from a separate, large genetic association study of Covid-19 outcomes is presented in the **Supplementary Appendix**.

Strengths of this trial included the randomized, double-blind, placebo-controlled design and stratification based on disease severity and corticosteroid use. In addition, unlike the RECOVERY trial, we systematically collected all SAEs and AESIs providing important safety information of the use of sarilumab of doses up to 800 mg in the hospitalized Covid-19 population.

Our trial also has several limitations. The phase 3 sample size estimation based on the phase 2 interim data may have led to an underpowered study, which was further impacted by the 2:1 randomization ratio, leading to a small placebo treatment group. The heterogeneity in the critically ill population as seen in clinical trials in acute respiratory distress syndrome not related to Covid-19 highlights the need for large simple trials such as RECOVERY and SOLIDARITY to adequately assess efficacy of therapeutics in the hospitalized Covid-19 population.^27,28^ Another limitation is that the 400-mg dose may be sub-therapeutic, as suggested by a reduction in CRP concentration in the first 7 days and a subsequent rebound (**Fig. S3**) thereafter, with similar findings reported in a separate trial using sarilumab 400 mg.^29^ Although an 800-mg treatment arm was included late in the trial, the small sample size makes interpretation challenging.

Based on results from the RECOVERY and REMAP-CAP trials, IL-6R inhibitors are currently recommended in combination with corticosteroids in certain hospitalized patients.^30,31^ Within the REMAP-CAP study, although the point estimates for tocilizumab and sarilumab are similar, the smaller sample size in the sarilumab group and the lack of benefit of sarilumab in multiple small trials leave open the question about whether the benefit demonstrated with tocilizumab in RECOVERY is a class effect.^32^ The recent addition of tocilizumab and sarilumab into treatment guidelines will make conducting large, controlled trials with sarilumab challenging.^30,31,33^ Therefore, to further inform decisions on the use of sarilumab, a meta-analysis of multiple clinical trials with sarilumab to account for heterogeneous patient populations, SOC protocols, and treatment effects will be critical and is currently being conducted.^34^

## Supporting information

MNgGong ICMJE form

MO'Brien ICMJE form

MSobieszczyk ICMJE form

NBraunstein ICMJE form

NHajizadeh ICMJE form

RBhore ICMJE form

RHosain ICMJE form

SBrown ICMJE form

SKSaggar ICMJE form

SSivplasingam ICMJE form

SSomersan-Karakaya ICMJE form

SSperber ICMJE form

TDiCoccio ICMJE form

VMenon ICMJE form

WKampman ICMJE form

WPark ICMJE form

ABaras ICMJE form

ABoyapati ICMJE form

AGiannelou ICMJE form

AMahmood ICMJE form

AWaldrom ICMJE form

BAkinlade ICMJE form

BMusser ICMJE form

DGulabani ICMJE form

DLederer ICMJE form

DRamesh ICMJE form

DStein ICJME form

DWeinreich ICMJE form

ELabriola-Tompkins ICMJE form

GCriner ICMJE form

GHerman ICMJE form

GYancopoulos ICMJE form

JAberg ICMJE form

JHorowitz ICMJE form

JKosmicki ICMJE form

JParrino ICMJE form

LPurcell ICMJE form

MCNivens ICMJE form

MFerreira ICMJE form

MFerreira ICMJE form

MFerreira ICMJE form

## Data Availability

Qualified researchers may request access to study documents (including the clinical study report, study protocol with any amendments, blank case report form, statistical analysis plan) that support the methods and findings reported in this manuscript. Individual anonymized participant data will be considered for sharing once the indication has been approved by a regulatory body, if there is legal authority to share the data and there is not a reasonable likelihood of participant re-identification. Submit requests to https://vivli.org/.

## ACKNOWLEDGMENT

The authors thank the patients, their families, and investigational site members involved in this study (principal investigators and subprincipal investigators listed in the Supplementary Appendix – Study Sites and Investigators); the members of the Independent Data Monitoring Committee (Steve Dahlberg, M.S., Mitchell Levy, M.D., Victor Ortega, M.D., Ph.D., Kevin Winthrop, M.D., Thomas Cook, Ph.D. [non-voting member], Caryn Trbovic, Ph.D., S. Balachandra Dass, Ph.D., and Brian Head, Ph.D., from Regeneron Pharmaceuticals) for assistance with development of the manuscript; and Prime, Knutsford, United Kingdom, for manuscript formatting and copy-editing suggestions.

We dedicate this article to the memory of Colby Burk, M.S., for his contributions and commitment to patients with Covid-19.

## DATA SHARING

A data sharing statement provided by the authors is available with the full text of this article.

