## Supplementary material for "A Randomized Placebo-Controlled Trial of Sarilumab in Hospitalized Patients with Covid-19": MNgGong ICMJE form

### SUPPLEMENTARY APPENDIX

### Study Sites and Investigators

| Name of Principal Investigator | Site Name, State |
|---|---|
| Judith Aberg | Mt. Sinai – Morningside, NY<br>Mt. Sinai – West, NY<br>Mt. Sinai – Beth Israel, NY<br>Mt. Sinai – Icahn School of Medicine, NY |
| Timothy Albertson | University of California Davis, CA |
| Danny Branstetter | Wellstar Kennestone, GA |
| Mark Brantly | University of Florida, FL |
| Daniel Brotman | Johns Hopkins University, MD |
| Samuel M. Brown | Intermountain Medical Center, UT |
| Thomas Campbell | University of Colorado, Denver, CO |
| Steven Chang | UCLA Division of Pulmonary and Critical Care, CA |
| Donald Chen | Westchester Medical Center, NY |
| Geoffrey Chupp | Yale New Haven Hospital, CT |
| Gerard J. Criner | Lewis Katz School of Medicine at Temple University, PA |
| Marjolein de Wit | Virginia Commonwealth, VA |
| George Diaz | Providence – Everett Regional Medical Center, WA |
| Svetolik Djurkovic | Inova Fairfax Medical Campus, VA |
| Christian Engell | Newark Beth Israel Medical Center, NJ |
| Jay Finigan | National Jewish Health – Denver, CO |
| Bettina Fries | Stony Brook University, NY |
| Dahlene Fusco | Tulane, LA |
| Marshall Glesby | Cornell Weill Medical Center, NY |
| Michelle Gong | Montefiore – Weiler Hospital, NY<br>Montefiore – Moses, NY |
| Robert Gottlieb | Baylor University Medical Center, TX |
| Kevin Gregg | University of Michigan, MI |
| Negin Hajizadeh | Northwell Health – Lenox Hill, NY<br>Northwell Health, Northshore University Hospital, Manhasset, NY<br>Northwell Health, Long Island Jewish Hospital, New Hyde Park, NY |
| Robert Hallowell | Beth Israel Deaconess Medical Center, MA |
| Teresa Hammond | Providence – St. Johns, CA |
| Akram Khan | Oregon Health & Science University, OR |
| Kami Kim | University of South Florida, FL |
| Andreas Klein | Tufts Medical Center, MA |
| Jerry Krishnan | University of Illinois at Chicago, IL |
| Yu Shia Lin | Maimonides Medical Center, NY |
| Edward Liu | Hackensack Meridian School of Medicine and Jersey Shore University Medical Center, NJ |
| George Marshall Lyon | Emory University Hospital, GA |
| Anuj Malik | Ascension St. John Hospital, OK |
| Jain Mamta | UT South Western, TX |
| Michael K. Mansour | Massachusetts General Hospital, MA |
| Vidya Menon | Lincoln Medical Center, NY |
| Amay Parikh | Florida Hospital, Orlando, AdventHealth, FL |
| William Park | UW Medicine/Valley Medical Center, WA |
| Igor Puzanov | Roswell Park – Buffalo, NY |
| Farbod Raiszadeh | Harlem Hospital, NY |
| Raymond Razonable | Mayo Clinic, MN |

| | |
|---|---|
| Fariborz Rezai | Saint Barnabas Medical Center – RWJBarnabas Health, NJ |
| Ruxana Sadikot | Atlanta VA Medical Center, GA |
| Suraj Saggar | Holy Name Medical Center, NJ |
| Carlos Salama | Elmhurst Hospital, NY |
| John Sensakovic | Hackensack Meridian School of Medicine and JFK Hospital, NJ |
| Paul Simonelli | Geisinger – Medical Center, PA<br>Geisinger – Community Medical Center, PA<br>Geisinger – Wyoming Valley, PA |
| Magdalena Sobieszczyk | Columbia University, NY |
| Steven Sperber | Hackensack Meridian School of Medicine and Hackensack University Medical Center – Hackensack, NJ |
| David Stein | Jacobi Medical Center, NY |
| Daniel Sterman | New York University, NY |
| Stephen Walsh | Brigham and Women’s Hospital, MA |
| Jason Wells | Providence Health and Services, Portland, OR |
| Eric Whitman | Atlantic Center for Research, NJ |
| Glenn Wortmann | MedStar (Washington Hospital Center), Washington, DC |
| Richard G. Wunderink | Northwestern University, IL |
| Daniel Kett | University of Miami, UMMG Infection Control, FL |

### Regeneron Sarilumab-COVID-19 Study Team

Moetaz Albizem, Gayatri Anand, Dhanalakshmi Barron, Georgia Bellingham, Alison Brown, Colby Burk, Wilson Caldwell, Michael N. Cantor, S. Balachandra Dass, John Davis, Ajla Dupljak, Haitao Gao, Evelyn Gasparino, Lori Geisler, Ruchin Gorawala, Ingeborg Heirman, Olga Herrera, Matthew Houghton, Susan Irvin, Denise Kennedy, Yasmin Khan, Carol Lee, Kelly Lewis-Amezcua, Qin Li, Leah Lipsich, Adnan Mahmood, Marco Mancini, Jutta Miller, Michael Partridge, Christina Perry, Valancia Reddick, Arsalan Shabbir, Nirav Shah, Kenneth Turner, Dana Wolken, Joseph Wolken, Sonia Yanes, and Jeannie Yo.

### Supplementary Methods

#### Inclusion and Exclusion Criteria

##### *Inclusion Criteria*

A patient must meet the following criteria to be eligible for inclusion in the study:

1) Male or female adult ≥18 years of age at time of enrollment
2) Hospitalized (or documentation of a plan to admit to the hospital if the patient is in an emergency department) with illness of any duration with evidence of pneumonia by chest radiograph, chest computed tomography or chest auscultation (rales, crackles), requires supplemental oxygen and/or assisted ventilation and meets 1 of the following criteria:

**Phase 2 and Phase 3 Cohort 1**: must meet at least 1 of the following at baseline (patients meeting more than one criterion will be categorized in the most severely affected category):

- **Severe disease**
  - Requires supplemental oxygen administration by nasal cannula, simple face mask, or other similar oxygen delivery device (i.e., above pre–COVID-19 baseline requirement, if any, by the patient)
- **Critical disease**
  - Requires supplemental oxygen delivered by non-rebreather mask or high-flow nasal cannula, <u>OR</u>
  - Use of invasive or non-invasive ventilation, <u>OR</u>
  - Requiring treatment in an intensive care unit

- **Multi-system organ dysfunction**
  - Multi-system organ dysfunction**:** use of vasopressors, extracorporeal life support, or renal replacement therapy

    Note: patients receiving vasopressors for reasons other than circulatory shock (e.g., related to sedation or mechanical ventilation) may be randomized into the critical stratum
- **Immunocompromised**
  - Immunocompromised patients (or on immunosuppressant treatments)

1. Laboratory-confirmed SARS-CoV-2 infection as determined by a polymerase chain reaction (PCR) result from any specimen (or other commercial or public health assay) within 2 weeks prior to randomization and no alternative explanation for current clinical condition
2. Willing and/or able to comply with study-related procedures/assessments
3. Provide informed consent signed by study patient or legally acceptable representative

***Exclusion Criteria***

A patient who meets any of the following criteria will be excluded from the study:

1. In the opinion of the investigator, not expected to survive for more than 48 hours from screening

2. Presence of any of the following abnormal laboratory values at screening: absolute neutrophil count <2000 $mm^3$, aspartate aminotransferase or alanine aminotransferase >5x upper limit of normal, platelets <50,000 per $mm^3$
3. Treatment with anti–IL-6, anti–IL-6R antagonists, or with Janus kinase inhibitors (JAKi) in the past 30 days or plans to receive during the study period
4. Current treatment with the simultaneous combination of leflunomide and methotrexate
5. Exclusion criteria related to tuberculosis (TB)
    a. Known active TB or a history of incompletely treated TB
    b. Suspected or known extrapulmonary TB
6. Patients with suspected or known active systemic bacterial or fungal infections

    Note: Patients with a history of positive bacterial or fungal cultures but on enrollment do not have suspected or known active systemic bacterial or fungal infections may be enrolled
7. Participation in a double-blind clinical research study evaluating an investigational product or therapy within 3 months and less than 5 half-lives of investigational product prior to the screening visit

    Exception: The use of remdesivir, hydroxychloroquine, or other treatments being used for COVID-19 treatments in the context of an open-label study, emergency use authorization, compassionate use protocol, or open-label use is permitted
8. Any physical examination findings, and/or history of any illness, concomitant medications, or recent live vaccines that, in the opinion of the study investigator,

might confound the results of the study or pose an additional risk to the patient by their participation in the study

9. Known systemic hypersensitivity to sarilumab or the excipients of the drug product

#### Dose Preparation

Sarilumab (Kevzara®) was provided in a 200 mg pre-filled syringe containing 175 mg/mL of active drug and was administered intravenously (IV) over 1-hour via bags containing 100 mL of 0.9% sodium chloride solution, followed by flush with 0.9% sodium chloride for an additional 15 minutes.

The infusion was stored at room temperature prior to administration and was administered within 4 hours of preparation. Infusion through either central or peripheral lines was allowed. Direct mixing of sarilumab with other drug(s) was prohibited. When it was needed to administer sarilumab concomitantly with other drug(s) (i.e., if a common IV infusion set or central catheter [central line] was used), appropriate steps (e.g., flush before and after administration of sarilumab) was recommended to prevent sarilumab from mixing with other drug(s) while in the infusion set or central catheter.

#### Definitions of Disease Severity Strata

Severe stratum was defined as patients receiving low-flow supplemental oxygen. Critical stratum was defined as requiring supplemental oxygen by nonrebreather mask or high-flow nasal device, noninvasive ventilation, invasive mechanical ventilation (MV), or management in an intensive care unit (ICU). Multi-system organ dysfunction (MSOD) stratum was defined as use of vasopressors, extracorporeal life support, or renal replacement therapy.

#### Study Drug Administration and Data Collection

Sarilumab 200 mg, 400 mg, or normal saline (used as placebo for sarilumab) were administered intravenously over 1 hour. An unmasked pharmacist was responsible for preparation of all study interventions. Patients, care providers, outcome assessors, and investigators remained masked to assigned intervention throughout the study. Study drug was administered on day 1 (baseline). Following protocol amendment 4 (April 6, 2020), patients were eligible to be re-dosed for clinical worsening at 24 hours and weekly for up to four doses. Data were collected daily from baseline until day 29 or hospital discharge, and at the end of the study (day 60).

#### Summary of Key Protocol Amendments and Study Adaptations

Key amendments to the study in the course of adapting the phase 3 design based on phase 2 results.

| Protocol Amendment (Date) | Key Study Design Update |
|---|---|
| **Initial Study Design** (March 15, 2020) | • Phase 2/3 study. Patients randomized 2:2:1 to sarilumab 400 mg IV, sarilumab 200 mg IV, or placebo<br>• Phase 2 primary end point: time to resolution of fever |
| **Protocol Amendment 1** (March 19, 2020) | • Aligned safety objectives and end points with adverse events of special interest<br>• Updated from a 6-point clinical status scale to a 7-point clinical status scale<br>• Updated inclusion criteria to accept a positive PCR result within 2 weeks of randomization<br>• Nasopharyngeal swab added as an acceptable method to collect virus based on emerging guidelines for SARS-CoV-2 testing |
| **Protocol Amendment 2** | • Withdrawn and not implemented |
| **Protocol Amendment 3** (March 28, 2020) | • Phase 2 primary end point changed to percent reduction in C-reactive protein (CRP)<br>• Reduced number of patients for phase 2<br>• Removed the requirement for a documented fever at randomization<br>• Removed exclusion criteria that previously prohibited patients who received immunosuppressive therapies from participating in this study |
| **Protocol Amendment 4** (April 6, 2020) | • Re-dosing permitted after 24 hours of dosing if no clinical response is observed<br>• Phase 2 sample size increased<br>• Removal of exclusion for immunocompromised patients |
| **Protocol Amendment 5** (May 9, 2020) | • Based on feedback from the Independent Data Monitoring Committee (IDMC), patients will no longer receive sarilumab 200 mg or be eligible for repeat dosing of 200 mg, and patients in the severe and Multi-System Organ Dysfunction (MSOD) stratum will no longer receive sarilumab 400 mg or placebo |

| | |
|---|---|
| | • Cohort 1 of phase 3 will complete enrollment after ~170 patients on a ventilator at baseline have been randomized in the critical stratum. Cohort 2 of phase 3, in which patients are randomized to receive sarilumab 800 mg or placebo, will start enrolling patients upon the completion of enrollment in cohort 1. |
| **Protocol Amendment 6** (May 16, 2020) | Based on regulatory authority feedback:<br>• As part of the phase 3 adaptation, two additional independently powered cohorts were added to evaluate sarilumab 800 mg versus placebo in patients on mechanical ventilation at baseline (cohort 2) and in patients not on mechanical ventilation but on high-intensity oxygen therapy without mechanical ventilation at baseline (cohort 3)<br>• Key secondary end point of all-cause mortality changed from<br>• Day 60 to day 29 |
| **Protocol Amendment 7** (June 11, 2020) | Based on feedback from the IDMC, the following changes were made to the protocol:<br>• In phase 3 cohort 1, only patients receiving mechanical ventilation will be re-dosed<br>• In phase 3 cohort 2, patients will only receive study drug while receiving mechanical ventilation<br>• In phase 3 cohort 3, no additional patients will be enrolled and patients on study will not be re-dosed |

#### Clinical Status Scale

The 7-point ordinal scale is an assessment of the clinical status. The scale is as follows:

- 1, Death
- 2, Hospitalized, requiring invasive mechanical ventilation or extracorporeal membrane oxygenation
- 3, Hospitalized, requiring non-invasive ventilation or high-flow oxygen devices
- 4, Hospitalized, requiring supplemental oxygen
- 5, Hospitalized, not requiring supplemental oxygen – requiring ongoing medical care (COVID-19–related or otherwise)
- 6, Hospitalized, not requiring supplemental oxygen – no longer requires ongoing medical care
- 7, Not hospitalized

#### Safety End Points

Safety outcomes included treatment emergent serious adverse events (SAEs) and the following adverse events of special interest (AESIs): grade 4 neutropenia (absolute neutrophil count $<500/\text{mm}^3$); confirmed invasive bacterial or fungal infections; elevations in alanine aminotransferase (ALT) or aspartate aminotransferase (AST) ≥3X ULN (for patients with normal baseline) or >3X ULN and at least 2-fold increase from baseline values (for patients with abnormal baseline); and grade 2 or higher hypersensitivity or infusion-related reactions.

#### Study Oversight

The protocol was developed by the sponsor (Regeneron Pharmaceuticals Inc.). Data were collected by the study investigators and analyzed by the sponsor. The local institutional review board or ethics committee at each study center oversaw trial conduct. All patients provided written informed consent before participating in the trial. The trial was overseen by an IDMC.

#### Ethics declaration

Ethics approval was obtained from the following ethics review boards: WCG IRB, Puyallup, WA (IRB00000533); Oregon Health & Science University, Portland, OR (MOD00027616); Institutional Review Board University of Florida, Gainesville, FL (IRB202000779); Columbia University, Human Research Protection Office and IRBs, New York, NY (IRB-AAAS9615); Johns Hopkins Medicine, Office of Human Subjects Research, IRB, Baltimore, MD (IRB00246576/CIR00058074); NYU School Of Medicine, New York, NY (I20-00351_MOD03); Northwestern University IRB, Chicago, IL (STU00212239-MOD0012); Westchester Medical Center, NY Medical College, Office of Research Administration, Valhalla, NY (IRBReg#00000428); Portland Health and Services IRB Providence – St. Johns, Portland, OR (MOD2020001024); Cornell Weill Medicine IRB, New York, NY (IRB00009419); Providence St. Joseph IRB, Renton, WA (MOD2020001029); Providence Health and Services IRB, Portland, OR (MOD2020000519); Ascension St. John Hospital, Tulsa, OK (IRB#00001980); Geisinger IRB, Danville, PA (IRB#0008345).

#### Statistical Methods

All analyses and tabulations were performed either using the Statistical Analysis Software (SAS) Version 9.4 or R language.

##### Adaptive Phase 2/3 Study Design

This randomized controlled clinical trial in hospitalized patients with COVID-19 across various disease severity stages followed a phase 2/3 adaptive design. Randomization was stratified by disease severity: 1) severe, 2) critical, 3) multi-system organ dysfunction (MSOD), and 4) immunocompromised, and by use of systemic corticosteroids. Due to the novel nature of COVID-19, efficacy end points at the time of study design were not well established. The phase 2/3 adaptive design of the study allowed for identification of early signs of clinical efficacy in the phase 2 data (e.g., reduction in C-reactive protein, and estimation of effects on clinical status measured on an ordinal scale). Interim phase 2 data analysis allowed adaptation of the phase 3 study design, including: 1) selection of clinical outcome end points in phase 3 (e.g., well-defined end points based on clinical status assessment of patients), 2) confirmation of the phase 3 population, 3) evaluation of further doses of sarilumab and 4) sample size adjustment for phase 3.

Although enrollment in the phase 2 and phase 3 portions of the study was seamless, the analyses of phase 2 and phase 3 portions of the study were kept separate. Each phase of the study was powered for separate objectives. No patients from phase 2 were included in the analysis of phase 3.

**Sample Size Calculations in Phase 3 Portion of the Study**

Based on the phase 2 interim data which included ~460 patients randomized (2:2:1) and stratified by disease severity and use of systemic corticosteroids manner to sarilumab 400 mg IV, sarilumab 200 mg IV, or placebo. Based on IDMC recommendations, the phase 3 design was adapted to allow continued enrollment of patients in the critical disease severity stratum only with the sarilumab 400-mg dose and placebo groups. Enrollment in other disease severity categories and in the sarilumab 200-mg group was halted.

In the phase 2 portion of the study, in a subgroup analysis of the critical stratum based on being on a mechanical ventilator at baseline, in the mechanical ventilator group, approximately 17% of patients treated with placebo (i.e., standard of care) and 57% of patients treated with sarilumab 400 mg (on top of standard of care) achieved ≥1-point improvement by day 22. Therefore, the sample size for phase 3 cohort 1 was recalculated using the chosen phase 2 end point of the proportion of patients with ≥1-point improvement in clinical status on the ordinal scale at day 22 in patients in the critical stratum who were on mechanical ventilation at baseline. With a 2:1 randomization ratio (sarilumab 400 mg IV:placebo), an effect size the same as that observed in phase 2, and 170 patients in the critical stratum on mechanical ventilation, the comparison between sarilumab 400 mg IV (n~113) and placebo (n~57) would have >99% power. Assuming the rate on placebo is 17% and the rate on sarilumab 400 mg is 37% (i.e., a difference in proportions is one-half that observed in phase 2), then the sample size of 170 would provide approximately 80% power to detect this reduced difference. These calculations assumed $\alpha=0.045$, allowing for an interim analysis at the 0.005 level. Therefore, this study planned to enroll approximately 450 critical patients to have approximately 170 patients within the critical stratum who are on mechanical ventilation and randomized to 400 mg IV or placebo.

Note that the total number of patients enrolled in phase 3 is larger than that to account for the patients who had been randomized to the severe stratum, MSOD stratum, or to the 200 mg IV dose group before the IDMC decision to discontinue further enrollment into those strata and dose groups. In addition, phase 3 adaptations also included two new cohorts of patients randomized 1:1 to sarilumab 800 mg IV (higher dose) or placebo, namely, cohort 2 consisting of patients who were on mechanical ventilation at baseline and cohort 3 consisting of patients not on mechanical ventilation but on high intensity oxygen therapy at baseline. These cohorts did not begin enrollment until after cohort 1 enrollment was completed and are not included in this analysis.

**<u>Analysis Populations</u>**

1) Intention-to-Treat (ITT) Population

The intention-to-treat (ITT) population included all randomized patients who received at least one dose of the study drug. Analysis of the ITT population was done according to the initial treatment assigned to the patient (as randomized).

The ITT population was the primary analysis population for phase 3. For phase 3, the specific primary population was the subset of patients in the critical stratum who were on mechanical ventilation (without extracorporeal membrane oxygenation [ECMO]) at baseline and randomized to sarilumab 400 mg or placebo. Demographics, baseline characteristics, and patient disposition was summarized in the ITT population.

2) Safety Population

The safety population (SAF) included all randomized patients who received at least one dose of the study drug. Analysis of the SAF population was done according to the treatment received (as treated). Determination of "as treated" will be based on the actual study drug received on day 1. The SAF population was used for analysis of all safety data, treatment exposure, medical history, and medication use.

**Analysis of Primary Efficacy Variable and Secondary Efficacy Variables**

Post adaptations to the phase 3 study design, the main focus of the efficacy analysis was pairwise comparison between sarilumab 400 mg and placebo in cohort 1 critical patients with mechanical ventilation without ECMO at baseline, in terms of the efficacy variable of proportion of patients with ≥1-point improvement in clinical status from baseline to day 22 using the 7-point ordinal scale. The primary analysis population was ITT.

Hypothesis test of superiority of sarilumab (400 mg) versus placebo was planned to be done using the stratified Cochran-Mantel-Haenszel (CMH) test for two proportions. Stratification factor was use of steroids at baseline (yes/no). Estimation of the treatment effect was provided as differences in proportions and confidence intervals calculated using the strata-adjusted confidence intervals from CMH method. P values and confidence intervals were reported with overall Type 1 error controlled at 0.05 (2-sided).

The statistical methods used for the key secondary efficacy variables were the same as the methods described for the primary efficacy variable.

These key secondary end points for the phase 3 cohort 1 portion of the study were to be tested sequentially in a hierarchical manner, while preserving the overall significance level at 0.05 (2-sided).

1. Proportion of patients with at least 1-point improvement in clinical status assessment from baseline to day 22 in patients with critical COVID-19
2. Proportion of patients who recover (discharged, or alive without supplemental oxygen use or at pre-COVID oxygen use) by day 22 in patients with critical COVID-19 receiving mechanical ventilation without ECMO at baseline
3. Proportion of patients who die through day 29 in patients with critical COVID-19 receiving mechanical ventilation without ECMO at baseline
4. Proportion of patients who die through day 29 in patients with critical COVID-19

Other secondary efficacy variables such as time-to-event end points were summarized with Kaplan-Meier estimates of medians and 95% confidence intervals on medians. Confidence intervals for hazard ratios comparing treatment groups were to be reported using Cox proportional hazards model with stratification factors mentioned earlier.

Cumulative incidence rates were plotted with comparisons between groups to be descriptively tested using log-rank test.

**Adjustment for Multiple Comparisons**

Analysis of phase 2 data was strictly separately done from phase 3. No patients from phase 2 were used in the phase 3 analyses. Multiplicity control in phase 3 for testing primary and key secondary end points using a hierarchical testing order was pre-specified in the Statistical Analysis Plan. Overall type 1 error was controlled within each cohort at 0.05 (2-sided) level.

### Human Genetic Studies Methods

#### Studies

We report association analyses from the COVID-19 Host Genetics Initiative (January 18, 2021, release, downloaded from https://www.covid19hg.org/results/).[1]

#### Definition of COVID-19 Outcomes

Risk of hospitalization was analyzed among RNA polymerase chain reaction (PCR)–confirmed SARS-CoV-2 positive cases (N=12,888) and controls who were either PCR-negative or not tested (N=1,295,966 population controls). Risk of severe COVID-19 was analyzed among individuals with a PCR-confirmed SARS-CoV-2 infection and record of mechanical ventilation or death due to COVID-19 (N=5,780) and population controls (N=1,115,203).

#### Association Analyses

The association between each phenotype and imputed variants was performed as described (https://www.covid19hg.org/results/r5/) with SAIGE v0.8[2] separately for each study and ancestry, with age, $age^2$, sex, age-by-sex, and 10 ancestry-informative principal components included as covariates. Association results were combined across studies and ancestries with an inverse-variance meta-analysis using METAL.[3]

### Phase 2 Study Results

#### Phase 2: Flow Diagram

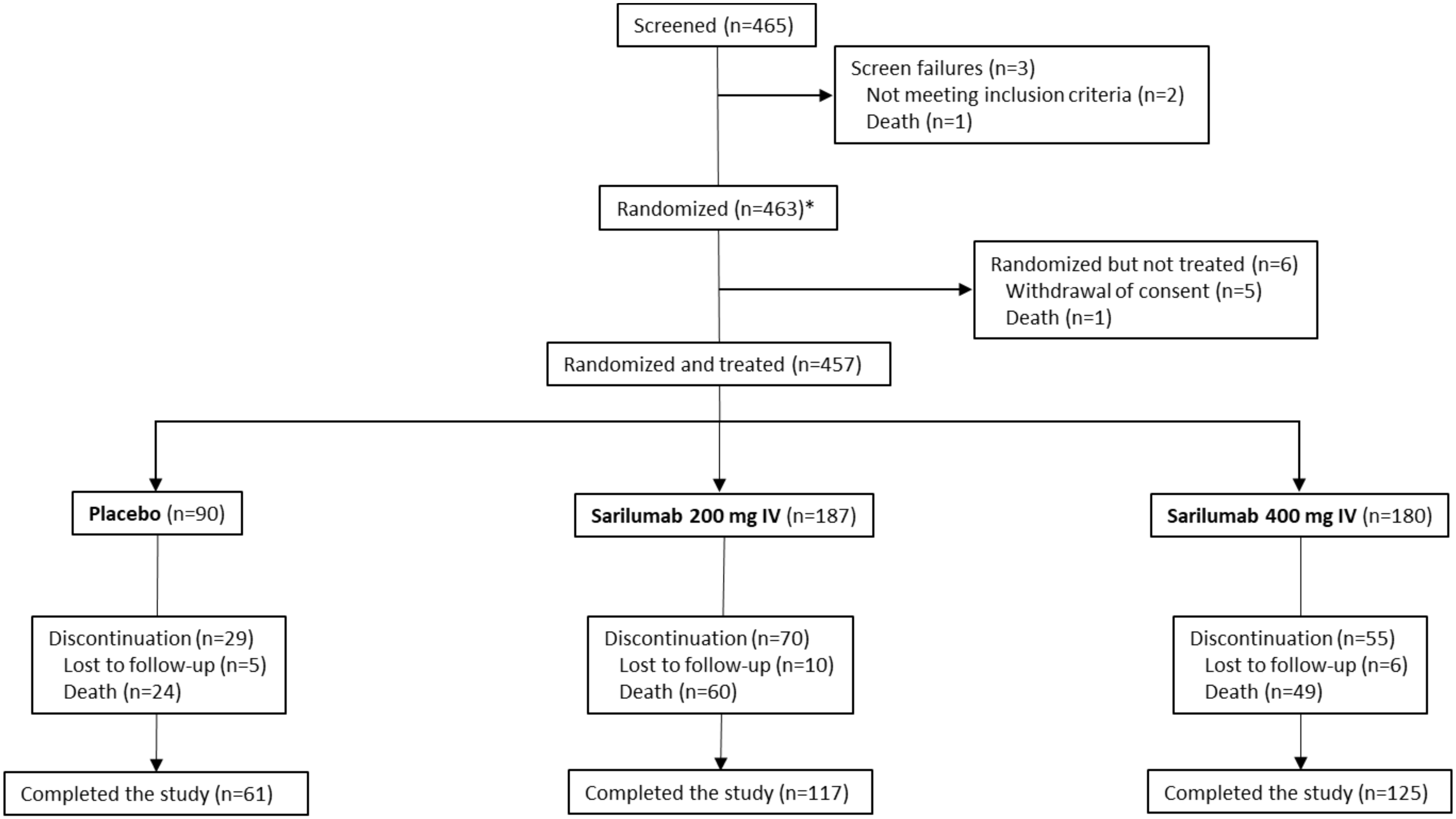

*Includes 1 screen failure that was randomized.

### Phase 2: Demographics and Baseline Characteristics (ITT Population)*

| | Severe Patients | | | | Critical Patients | | | | MSOD/Immunocompromised Patients | | | |
|---|---|---|---|---|---|---|---|---|---|---|---|---|
| | Placebo Group (N=25) | Sarilumab 200 mg IV Group (N=50) | Sarilumab 400 mg IV Group (N=51) | Total (N=126) | Placebo Group (N=44) | Sarilumab 200 mg IV Group (N=94) | Sarilumab 400 mg IV Group (N=88) | Total (N=226) | Placebo Group (N=21) | Sarilumab 200 mg IV Group (N=43) | Sarilumab 400 mg IV Group (N=41) | Total (N=105) |
| Median age— years (Q1:Q3) | 61.0 (55.0:68.0) | 56.5 (42.0:69.0) | 55.0 (43.0:68.0) | 57.0 (46.0:68.0) | 59.0 (48.5:67.5) | 61.0 (50.0:66.0) | 54.5 (43.5:65.5) | 58.5 (46.0:66.0) | 71.0 (53.0:73.0) | 60.0 (48.0:66.0) | 64.0 (49.0:70.0) | 61.0 (50.0:72.0) |
| Male sex — no. (%) | 15 (60.0) | 34 (68.0) | 38 (74.5) | 87 (69.0) | 37 (84.1) | 62 (66.0) | 70 (79.5) | 169 (74.8) | 17 (81.0) | 28 (65.1) | 30 (73.2) | 75 (71.4) |
| Ethnicity — no. (%) | | | | | | | | | | | | |
| Hispanic or Latino | 4 (16.0) | 13 (26.0) | 9 (17.6) | 26 (20.6) | 7 (15.9) | 29 (30.9) | 20 (22.7) | 56 (24.8) | 5 (23.8) | 12 (27.9) | 11 (26.8) | 28 (26.7) |
| Not Hispanic or Latino | 16 (64.0) | 29 (58.0) | 34 (66.7) | 79 (62.7) | 30 (68.2) | 44 (46.8) | 49 (55.7) | 123 (54.4) | 13 (61.9) | 25 (58.1) | 22 (53.7) | 60 (57.1) |
| Not reported | 4 (16.0) | 3 (6.0) | 7 (13.7) | 14 (11.1) | 4 (9.1) | 8 (8.5) | 8 (9.1) | 20 (8.8) | 2 (9.5) | 2 (4.7) | 2 (4.9) | 6 (5.7) |
| Unknown | 1 (4.0) | 5 (10.0) | 1 (2.0) | 7 (5.6) | 3 (6.8) | 13 (13.8) | 11 (12.5) | 27 (11.9) | 1 (4.8) | 4 (9.3) | 6 (14.6) | 11 (10.5) |
| Race — no. (%) | | | | | | | | | | | | |
| White | 15 (60.0) | 22 (44.0) | 22 (43.1) | 59 (46.8) | 16 (36.4) | 37 (39.4) | 29 (33.0) | 82 (36.3) | 8 (38.1) | 11 (25.6) | 13 (31.7) | 32 (30.5) |
| Black or African American | 4 (16.0) | 8 (16.0) | 8 (15.7) | 20 (15.9) | 6 (13.6) | 14 (14.9) | 16 (18.2) | 36 (15.9) | 5 (23.8) | 10 (23.3) | 7 (17.1) | 22 (21.0) |
| Asian | 1 (4.0) | 7 (14.0) | 6 (11.8) | 14 (11.1) | 6 (13.6) | 9 (9.6) | 10 (11.4) | 25 (11.1) | 2 (9.5) | 4 (9.3) | 5 (12.2) | 11 (10.5) |
| American Indian or Alaska Native | 0 | 0 | 0 | 0 | 0 | 0 | 0 | 0 | 0 | 0 | 0 | 0 |
| Native Hawaiian or Pacific Islander | 0 | 0 | 1 (2.0) | 1 (0.8) | 0 | 0 | 0 | 0 | 0 | 0 | 0 | 0 |
| Not reported | 3 (12.0) | 8 (16.0) | 7 (13.7) | 18 (14.3) | 9 (20.5) | 22 (23.4) | 24 (27.3) | 55 (24.3) | 3 (14.3) | 13 (30.2) | 13 (31.7) | 29 (27.6) |
| Other | 2 (8.0) | 5 (10.0) | 7 (13.7) | 14 (11.1) | 7 (15.9) | 12 (12.8) | 9 (10.2) | 28 (12.4) | 3 (14.3) | 5 (11.6) | 3 (7.3) | 11 (10.5) |
| Mean weight — kg | 86.63±21.14 | 91.32±22.14 | 89.97±24.53 | 89.81±22.82 | 93.59±20.73 | 89.96±22.10 | 90.79±18.88 | 90.97±20.56 | 88.50±18.21 | 96.09±31.29 | 90.38±22.92 | 92.32±25.88 |
| Body-mass index† | 29.93±6.12 | 31.36±7.82 | 30.58±7.90 | 30.77±7.50 | 31.37±6.83 | 31.76±7.91 | 30.78±5.47 | 31.30±6.80 | 30.14±5.30 | 33.74±8.88 | 32.94±12.67 | 32.65±9.95 |
| CRP, mg/l, n‡, median (min:max) | 25, 122.000 (5.10:335.23) | 48, 105.190 (14.70:296.70) | 49, 111.230 (1.00:517.01) | 122, 111.816 (1.00:517.01) | 42, 195.700 (11.10:603.01) | 92, 182.252 (5.00:819.94) | 84, 179.902 (8.68:662.00) | 218, 182.902 (5.00:819.94) | 20, 188.502 (52.50:564.81) | 40, 245.967 (37.50:654.66) | 38, 249.252 (28.00:589.71) | 98, 229.100 (28.00:654.66) |
| IL-6, pg/ml, n‡, median (min:max) | 22, 75.265 (9.59:328.77) | 43, 53.350 (9.10:323.83) | 45, 71.480 (9.10:2771.49) | 110, 67.105 (9.10:2771.49) | 42, 118.705 (9.10:2176.98) | 87, 127.530 (9.10:1597.06) | 82, 141.190 (9.10:21545.37) | 211, 131.900 (9.10:21545.37) | 18, 338.160 (23.89:2193.05) | 37, 208.800 (9.10:4058.18) | 33, 301.180 (23.29:9900.13) | 88, 254.945 (9.10:9900.13) |
| Comorbidities — no. (%) | | | | | | | | | | | | |
| Hypertension | 9 (36.0) | 20 (40.0) | 26 (51.0) | 55 (43.7) | 24 (54.5) | 38 (40.4) | 36 (40.9) | 98 (43.4) | 14 (66.7) | 20 (46.5) | 25 (61.0) | 59 (56.2) |
| Diabetes | 5 (20.0) | 5 (10.0) | 7 (13.7) | 17 (13.5) | 13 (29.5) | 22 (23.4) | 12 (13.6) | 47 (20.8) | 6 (28.6) | 14 (32.6) | 14 (34.1) | 34 (32.4) |
| Obesity§ | 9 (36.0) | 23 (46.0) | 18 (35.3) | 50 (39.7) | 20 (45.5) | 41 (43.6) | 41 (46.6) | 102 (45.1) | 10 (47.6) | 25 (58.1) | 17 (41.5) | 52 (49.5) |
| Systemic corticosteroid use — no. (%) | 3 (12.0) | 6 (12.0) | 6 (11.8) | 15 (11.9) | 7 (15.9) | 21 (22.3) | 16 (18.2) | 44 (19.5) | 4 (19.0) | 7 (16.3) | 6 (14.6) | 17 (16.2) |

*Plus–minus values are means ±SD. CRP denotes C-reactive protein, IL-6 interleukin-6, IV intravenous, MSOD multi-system organ dysfunction, SD standard deviation, Q quartile.
†The body-mass index is the weight in kilograms divided by the square of the height in meters.
‡Patients with available data.
§BMI >30 kg/m$^2$.

#### Phase 2: Primary Efficacy End Point (mITT population)*†

| Percent Change From Baseline to day 4 in CRP Levels | | | |
|---|---|---|---|
| | **Placebo Group (N=80)** | **Sarilumab 200 mg IV Group (N=152)** | **Sarilumab 400 mg IV Group (N=153)** |
| Mean change from baseline — log scale | -0.18±0.78 | -1.41±0.72 | -1.47±0.93 |
| LS mean difference vs. placebo — log scale | - | -1.25 | -1.30 |
| 95% CI | - | -1.456 to -1.035 | -1.511 to -1.090 |
| P value | - | <0.0001 | <0.0001 |
| LS mean percent reduction from baseline — raw scale | 20.0% | 77.0% | 78.0% |
| 95% CI | 3.5%–34.0% | 73.2%–80.3% | 74.6%–81.3% |

*Phase 2 mITT population includes all randomized patients who received at least one dose of the study drug and have high baseline IL-6 levels (above upper limit of normal). Plus–minus values are means ±SD. CI denotes confidence interval, IV intravenous, LS least-squares, mITT modified intention-to-treat, SD standard deviation.

†All disease severities.

### Phase 2: Summary of Selected Secondary End Points (ITT Population)

A. “Time to event” end points

| Outcome | Severe Patients | | Critical Patients | | MSOD/Immunocompromised Patients | |
|---|---|---|---|---|---|---|
| | Sarilumab 200 mg IV Group (N=50) HR (95% CI) | Sarilumab 400 mg IV Group (N=51) HR (95% CI) | Sarilumab 200 mg IV Group (N=94) HR (95% CI) | Sarilumab 400 mg IV Group (N=88) HR (95% CI) | Sarilumab 200 mg IV Group (N=43) HR (95% CI) | Sarilumab 400 mg IV Group (N=41) HR (95% CI) |
| Time to improvement (≥2 points) in clinical status | 1.17 (0.71– 1.93) | 0.74 (0.44–1.25) | 1.24 (0.73–2.08) | 1.64 (0.98–2.74) | 0.91 (0.42–1.97) | 0.90 (0.42–1.95) |
| Time to improvement (≥1 point) in clinical status | 1.16 (0.70–1.92) | 0.77 (0.46–1.30) | 1.14 (0.68–1.90) | 1.55 (0.94–2.56) | 0.96 (0.44–2.06) | 0.92 (0.43–1.98) |
| Time to resolution of fever for 48 hours without antipyretics or until discharge, in febrile patients at baseline | 1.27 (0.66–2.43) | 0.74 (0.39–1.41) | 1.61 (0.76–3.40) | 2.10 (1.01–4.40) | 0.98 (0.32–3.05) | 0.83 (0.25–2.71) |
| Time to improvement in oxygenation (increase in $SpO_2/FiO_2$ of 50 or greater) for at least 48 hours or until discharge | 0.87 (0.52–1.47) | 0.50 (0.29–0.88) | 0.76 (0.48–1.21) | 1.09 (0.69–1.72) | 0.81 (0.40–1.66) | 1.01 (0.50–2.02) |
| Time to saturation ≥94% on room air | 0.32 (0.11–0.94) | 0.38 (0.14–1.05) | 1.22 (0.70–2.14) | 0.83 (0.46–1.51) | 0.74 (0.36–1.52) | 0.71 (0.35–1.47) |
| Time to discharge or NEWS2 ≤2 for 24 hours | 0.97 (0.59–1.59) | 0.65 (0.39–1.09) | 1.23 (0.73–2.09) | 1.91 (1.14–3.20) | 0.97 (0.43–2.17) | 0.94 (0.42–2.10) |
| Time to all-cause mortality | NA* | NA* | 1.21 (0.66–2.21) | 0.70 (0.37–1.35) | 1.15 (0.52–2.53) | 1.02 (0.45–2.30) |
| Time to discharge | 1.18 (0.71–1.95) | 0.74 (0.44–1.25) | 1.12 (0.65–1.95) | 1.51 (0.88–2.59) | 0.92 (0.39–2.20) | 0.56 (0.22–1.45) |

*Hazard ratio could not be calculated, as the number of events was too small.
Blue = key secondary end point.

#### B. Categorical end points.*

| | Severe Patients | | | | | Critical Patients | | | | | MSOD/Immunocompromised Patients | | | | |
|---|---|---|---|---|---|---|---|---|---|---|---|---|---|---|---|
| Outcome | Placebo Group (N=25) no. (%) [95% CI] | Sarilumab 200 mg IV Group (N=50) no. (%) [95% CI] | Sarilumab 400 mg IV Group (N=51) no. (%) [95% CI] | Risk difference sarilumab 200 mg IV vs placebo [95% CI] | Risk difference sarilumab 400 mg IV vs placebo [95% CI] | Placebo Group (N=44) no. (%) [95% CI] | Sarilumab 200 mg IV Group (N=94) no. (%) [95% CI] | Sarilumab 400 mg IV Group (N=88) no. (%) [95% CI] | Risk difference sarilumab 200 mg IV vs placebo [95% CI] | Risk difference sarilumab 400 mg IV vs placebo [95% CI] | Placebo (N=21) no. (%) [95% CI] | Sarilumab 200 mg IV Group (N=43) no. (%) [95% CI] | Sarilumab 400 mg IV Group (N=41) no. (%) [95% CI] | Risk difference sarilumab 200 mg IV vs placebo [95% CI] | Risk difference sarilumab 400 mg IV vs placebo [95% CI] |
| Proportion with 1-point improvement (day 22) | 23 (92.0%) [81.4–100.0] | 46 (92.0%) [84.5–99.5] | 35 (68.6%) [55.9–81.4] | 0.0% [-13.0–13.0] | -23.4% [-40.0–-6.8] | 15 (34.1%) [20.1–48.1] | 45 (47.9%) [37.8–58.0] | 53 (60.2%) [50.0–70.5] | 13.8% [-3.5–31.0] | 26.1% [8.8–43.5] | 10 (47.6%) [26.3–69.0] | 17 (39.5%) [24.9–54.1] | 18 (43.9%) [28.7–59.1] | -8.1% [-34.0–17.8] | -3.7% [-29.9–22.5] |
| Proportion who recover (day 22) | 23 (92.0%) [81.4–100.0] | 46 (92.0%) [84.5–99.5] | 35 (68.6%) [55.9–81.4] | 0.0% [-13.0–13.0] | -23.4% [-40.0–-6.8] | 15 (34.1%) [20.1–48.1] | 39 (41.5%) [31.5–51.4] | 46 (52.3%) [41.8–62.7] | 7.4% [-9.8–24.6] | 18.2% [0.7–35.6] | 7 (33.3%) [13.2–53.5] | 13 (30.2%) [16.5–44.0] | 10 (24.4%) [11.2–37.5] | -3.1% [-27.5–21.3] | -8.9% [-33.0–15.1] |

IV denotes intravenous.

#### Phase 2: Overview of Adverse Events (Safety Population)*

| | Severe Patients | | | Critical Patients | | | MSOD/Immunocompromised Patients | | |
|---|---|---|---|---|---|---|---|---|---|
| | **Placebo Group (N=25)** | **Sarilumab 200 mg IV Group (N=50)** | **Sarilumab 400 mg IV Group (N=51)** | **Placebo Group (N=44)** | **Sarilumab 200 mg IV Group (N=94)** | **Sarilumab 400 mg IV Group (N=88)** | **Placebo Group (N=21)** | **Sarilumab 200 mg IV Group (N=43)** | **Sarilumab 400 mg IV Group (N=41)** |
| TEAEs – total no. | 11 | 32 | 47 | 81 | 145 | 141 | 40 | 84 | 83 |
| SAEs – total no. | 4 | 7 | 27 | 62 | 95 | 96 | 24 | 57 | 49 |
| AESIs – total no. | 7 | 27 | 26 | 34 | 61 | 53 | 17 | 37 | 46 |
| Serious AESIs – total no. | 0 | 2 | 6 | 15 | 11 | 8 | 1 | 10 | 12 |
| Patients with any TEAE – no. (%) | 7 (28.0) | 19 (38.0) | 25 (49.0) | 28 (63.6) | 69 (73.4) | 56 (63.6) | 16 (76.2) | 35 (81.4) | 33 (80.5) |
| Patients with any SAE – no. (%) | 1 (4.0) | 5 (10.0) | 16 (31.4) | 26 (59.1) | 56 (59.6) | 41 (46.6) | 12 (57.1) | 29 (67.4) | 27 (65.9) |
| Patients with any AESI – no. (%) | 7 (28.0) | 17 (34.0) | 16 (31.4) | 17 (38.6) | 38 (40.4) | 37 (42.0) | 11 (52.4) | 22 (51.2) | 23 (56.1) |
| Patients with any serious AESI – no. (%) | 0 | 2 (4.0) | 4 (7.8) | 10 (22.7) | 8 (8.5) | 7 (8.0) | 1 (4.8) | 8 (18.6) | 8 (19.5) |
| Patients with any TEAE leading to death – no. (%) | 0 | 2 (4.0) | 8 (15.7) | 14 (31.8) | 37 (39.4) | 22 (25.0) | 9 (42.9) | 20 (46.5) | 15 (36.6) |
| Patients with any TEAE leading to withdrawal from the study drug – no. (%) | 0 | 0 | 0 | 0 | 0 | 0 | 0 | 0 | 1 (2.4) |
| Patients with any TEAE leading to study infusion interruption – no. (%) | 0 | 0 | 0 | 0 | 1 (1.1) | 1 (1.1) | 0 | 0 | 0 |
| Related TEAEs – total no. | 2 | 17 | 14 | 5 | 20 | 30 | 6 | 18 | 22 |
| Related SAEs – total no. | 0 | 2 | 3 | 0 | 2 | 8 | 0 | 5 | 7 |
| Related AESIs – total no. | 2 | 17 | 14 | 5 | 20 | 25 | 6 | 17 | 21 |
| Related serious AESIs – total no. | 0 | 2 | 3 | 0 | 2 | 3 | 0 | 4 | 6 |
| Patients with related TEAE – no. (%) | 2 (8.0) | 13 (26.0) | 9 (17.6) | 4 (9.1) | 13 (13.8) | 19 (21.6) | 5 (23.8) | 11 (25.6) | 11 (26.8) |
| Patients with related SAE – no. (%) | 0 | 2 (4.0) | 2 (3.9) | 0 | 1 (1.1) | 7 (8.0) | 0 | 4 (9.3) | 4 (9.8) |
| Patients with related AESI – no. (%) | 2 (8.0) | 13 (26.0) | 9 (17.6) | 4 (9.1) | 13 (13.8) | 18 (20.5) | 5 (23.8) | 10 (23.3) | 10 (24.4) |
| Patients with related serious AESI – no. (%) | 0 | 2 (4.0) | 2 (3.9) | 0 | 1 (1.1) | 3 (3.4) | 0 | 3 (7.0) | 3 (7.3) |

*AESI denotes adverse event of special interest, IV intravenous, TEAE treatment-emergent adverse event, SAE serious adverse event.

#### Phase 2: Treatment-Emergent Serious Adverse Events and Adverse Events of Special Interest Occurring in ≥5% of Patients in Any Group (Safety Population)*

| System Organ Class<br>Preferred Term | Severe Patients | | | Critical Patients | | | MSOD/Immunocompromised Patients | | |
|---|---|---|---|---|---|---|---|---|---|
| | Placebo Group (N=25) | Sarilumab 200 mg IV Group (N=50) | Sarilumab 400 mg IV Group (N=51) | Placebo Group (N=44) | Sarilumab 200 mg IV Group (N=94) | Sarilumab 400 mg IV Group (N=88) | Placebo Group (N=21) | Sarilumab 200 mg IV Group (N=43) | Sarilumab 400 mg IV Group (N=41) |
| **SAEs** | | | | | | | | | |
| SAEs — no. | 4 | 7 | 27 | 62 | 95 | 96 | 24 | 57 | 49 |
| Patients with ≥1 SAE — no. (%) | 1 (4.0) | 5 (10.0) | 16 (31.4) | 26 (59.1) | 56 (59.6) | 41 (46.6) | 12 (57.1) | 29 (67.4) | 27 (65.9) |
| Vascular disorders — no. (%) | | | | | | | | | |
| Deep vein thrombosis | 1 (4.0) | 0 (0) | 2 (3.9) | 0 (0.0) | 3 (3.2) | 5 (5.7) | 0 (0.0) | 1 (2.3) | 2 (4.9) |
| Cardiac disorders — no. (%) | | | | | | | | | |
| Cardiac arrest | 0 (0.0) | 0 (0.0) | 2 (3.9) | 0 (0.0) | 6 (6.4) | 8 (9.1) | 3 (14.3) | 3 (7.0) | 6 (14.6) |
| Cardio-respiratory arrest | 0 (0.0) | 1 (2.0) | 2 (3.9) | 4 (9.1) | 9 (9.6) | 4 (4.5) | 2 (9.5) | 1 (2.3) | 2 (4.9) |
| Infections and infestations — no. (%) | | | | | | | | | |
| Staphylococcal infection | 0 (0.0) | 0 (0.0) | 0 (0.0) | 3 (6.8) | 0 (0.0) | 2 (2.3) | 0 (0.0) | 0 (0.0) | 2 (4.9) |
| COVID-19 | 0 (0.0) | 0 (0.0) | 1 (2.0) | 2 (4.5) | 7 (7.4) | 1 (1.1) | 1 (4.8) | 6 (14.0) | 3 (7.3) |
| Investigations — no. (%) | | | | | | | | | |
| Transaminases increased | 0 (0.0) | 0 (0.0) | 0 (0.0) | 1 (2.3) | 2 (2.1) | 1 (1.1) | 0 (0.0) | 4 (9.3) | 0 (0.0) |
| Respiratory, thoracic, and mediastinal disorders — no. (%) | | | | | | | | | |
| Respiratory failure | 0 (0.0) | 1 (2.0) | 6 (11.8) | 5 (11.4) | 11 (11.7) | 4 (4.5) | 1 (4.8) | 3 (7.0) | 2 (4.9) |
| Acute respiratory failure | 0 (0.0) | 0 (0.0) | 0 (0.0) | 6 (13.6) | 7 (7.4) | 3 (3.4) | 1 (4.8) | 1 (2.3) | 1 (2.4) |
| Renal and urinary disorders — no. (%) | | | | | | | | | |
| Acute kidney injury | 0 (0.0) | 1 (2.0) | 0 (0.0) | 5 (11.4) | 4 (4.3) | 6 (6.8) | 0 (0.0) | 4 (9.3) | 3 (7.3) |
| Renal failure | 0 (0.0) | 0 (0.0) | 0 (0.0) | 1 (2.3) | 1 (1.1) | 2 (2.3) | 0 (0.0) | 3 (7.0) | 0 (0.0) |
| **AESIs** | | | | | | | | | |
| AESIs — no. | 7 | 27 | 26 | 34 | 61 | 53 | 17 | 37 | 46 |
| Patients with ≥1 AESI — no. (%) | 7 (28.0) | 17 (34.0) | 16 (31.4) | 17 (38.6) | 38 (40.4) | 37 (42.0) | 11 (52.4) | 22 (51.2) | 23 (56.1) |
| Investigations — no. (%) | | | | | | | | | |
| Alanine aminotransferase increased | 2 (8.0) | 14 (28.0) | 10 (19.6) | 9 (20.5) | 18 (19.1) | 19 (21.6) | 4 (19.0) | 8 (18.6) | 4 (9.8) |
| Aspartate aminotransferase increased | 2 (8.0) | 7 (14.0) | 7 (13.7) | 7 (15.9) | 14 (14.9) | 14 (15.9) | 6 (28.6) | 11 (25.6) | 12 (29.3) |
| Transaminases increased | 1 (4.0) | 1 (2.0) | 3 (5.9) | 1 (2.3) | 6 (6.4) | 4 (4.5) | 1 (4.8) | 4 (9.3) | 0 (0.0) |
| Infections and infestations — no. (%) | | | | | | | | | |
| Enterococcal bacteremia | 0 (0.0) | 0 (0.0) | 0 (0.0) | 0 (0.0) | 0 (0.0) | 0 (0.0) | 0 | 0 | 3 (7.3) |
| Pneumonia | 0 (0.0) | 0 (0.0) | 0 (0.0) | 0 (0.0) | 2 (2.1) | 1 (1.1) | 1 (4.8) | 0 | 3 (7.3) |
| Staphylococcal bacteremia | 0 (0.0) | 0 (0.0) | 0 (0.0) | 0 (0.0) | 0 (0.0) | 0 (0.0) | 1 (4.8) | 0 | 3 (7.3) |
| Staphylococcal infection | 0 (0.0) | 0 (0.0) | 0 (0.0) | 4 (9.1) | 0 | 2 (2.3) | 1 (4.8) | 0 | 3 (7.3) |

*AESI denotes adverse event of special interest, IV intravenous SAE serious adverse event.

### Phase 3 (Cohort 2) Study Results

#### Phase 3 (Cohort 2): Flow Diagram

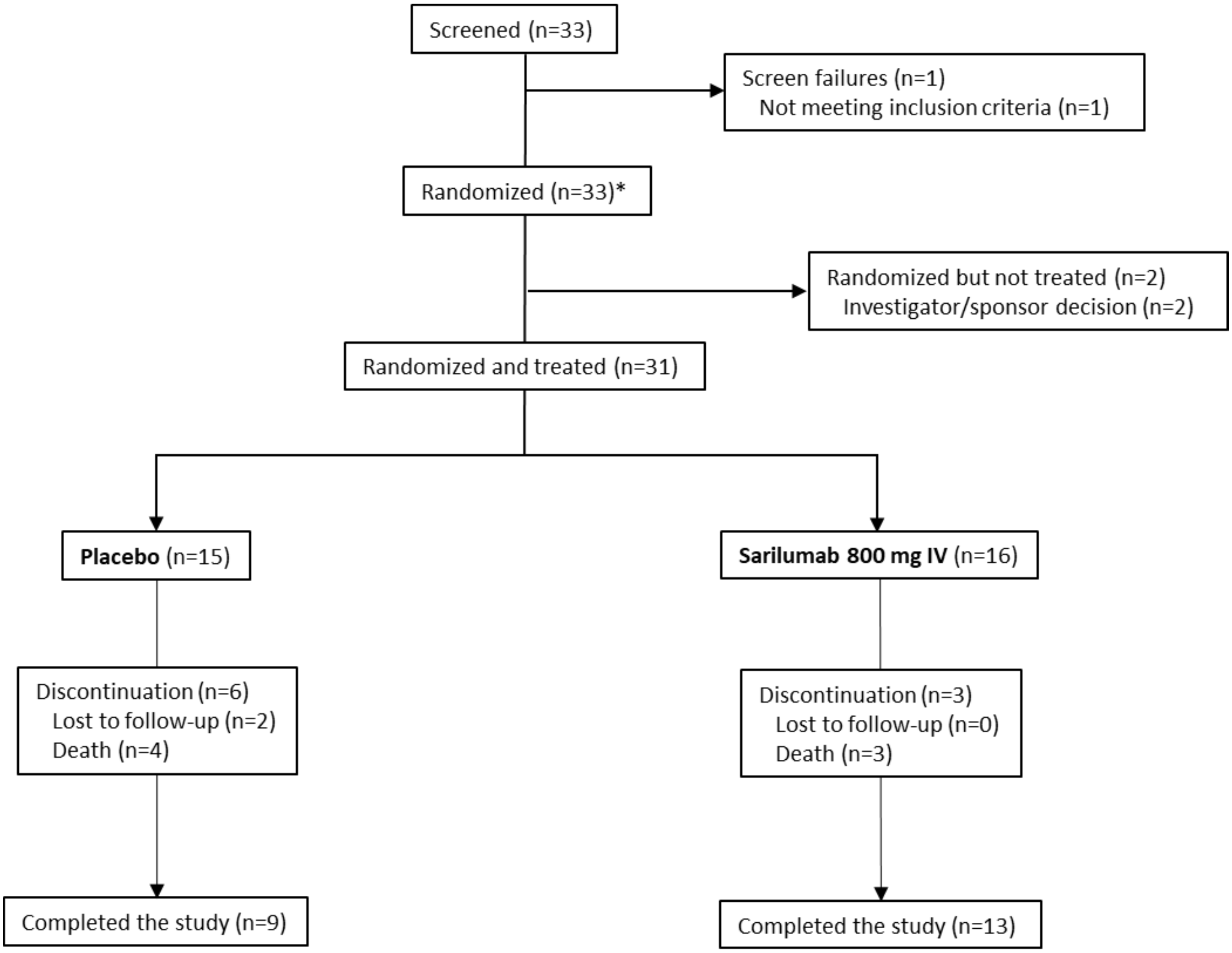

*Includes 1 screen failure that was randomized.

#### Phase 3 (Cohort 2): Demographics and Baseline Characteristics (ITT Population)*

| | Placebo Group (N=15) | Sarilumab 800 mg IV Group (N=16) |
|---|---|---|
| Median age — years (Q1:Q3) | 49 (41.0:58.0) | 48.5 (41.5:65.0) |
| Male sex — no. (%) | 11 (73.3) | 11 (68.8) |
| Ethnicity — no. (%) | | |
| Hispanic or Latino | 5 (33.3) | 10 (62.5) |
| Not Hispanic or Latino | 5 (33.3) | 6 (37.5) |
| Not reported | 1 (6.7) | 0 |
| Unknown | 4 (26.7) | 0 |
| Race — no. (%) | | |
| White | 7 (46.7) | 8 (50.0) |
| Black or African American | 3 (20.0) | 3 (18.8) |
| Asian | 0 | 0 |
| American Indian or Alaska Native | 0 | 0 |
| Native Hawaiian or Pacific Islander | 0 | 0 |
| Not reported | 5 (33.3) | 1 (6.3) |
| Other | 0 | 4 (25.0) |
| Mean weight – kg | 97.72±25.01 | 104.23±31.94 |
| Body-mass index† | 33.83±8.15 | 50.71±58.82 |
| CRP, mg/L, n‡, median (min:max) | 13, 185.904 (91.70:445.01) | 15, 184.000 (64.00:405.00) |
| IL-6, pg/mL, n‡, median (min:max) | 14, 115.475 (9.59:882.96) | 15, 164.730 (9.59:1689.34) |
| Comorbidities — no. (%) | | |
| Hypertension | 5 (33.3) | 8 (50.0) |
| Diabetes | 2 (13.3) | 7 (43.8) |
| Obesity§ | 10 (66.7) | 12 (75.0) |
| Systemic corticosteroid use — no. (%) | 10 (66.7) | 7 (43.8) |

*Plus–minus values are means ±SD. CRP denotes C-reactive protein, ITT intention-to-treat, IV intravenous, SD standard deviation.

†The body-mass index is the weight in kilograms divided by the square of the height in meters.

‡Patients with available data.

§Body-mass index >30 kg/$m^2$.

#### Phase 3 (Cohort 2): Overview of Adverse Events (Safety Population)*

| | **Placebo Group (N=15)** | **Sarilumab 800 mg IV Group (N=16)** |
|---|---|---|
| TEAEs — no. | 28 | 20 |
| SAEs — no. | 21 | 12 |
| AESIs — no. | 11 | 11 |
| Serious AESIs — no. | 4 | 3 |
| Patients with any TEAE — no. (%) | 11 (73.3) | 9 (56.3) |
| Patients with any SAE — no. (%) | 10 (66.7) | 5 (31.3) |
| Patients with any AESI — no. (%) | 7 (46.7) | 8 (50.0) |
| Patients with any serious AESI — no. (%) | 3 (20.0) | 2 (12.5) |
| Patients with any TEAE leading to death — no. (%) | 3 (20.0) | 2 (12.5) |
| Patients with any TEAE leading to withdrawal from the study drug — no. (%) | 0 | 1 (6.3) |
| Patients with any TEAE leading to study infusion interruption — no. (%) | 1 (6.7) | 1 (6.3) |
| Related TEAEs — no. | 9 | 7 |
| Related SAEs — no. | 4 | 2 |
| Related AESIs — no. | 8 | 7 |
| Related serious AESIs — no. | 3 | 2 |
| Patients with related TEAE — no. (%) | 5 (33.3) | 6 (37.5) |
| Patients with related SAE — no. (%) | 3 (20.0) | 1 (6.3) |
| Patients with related AESI — no. (%) | 5 (33.3) | 6 (37.5) |
| Patients with related serious AESI — no. (%) | 2 (13.3) | 1 (6.3) |

*AESI denotes adverse event of special interest, IV intravenous, TEAE treatment-emergent adverse event, SAE serious adverse event.

#### Phase 3 (Cohort 2): Treatment-Emergent Serious Adverse Events and Adverse Events of Special Interest Occurring in ≥5% of Patients in Any Group (Safety Population)*

| System Organ Class<br>Preferred Term | Placebo Group (N=15) | Sarilumab 800 mg IV Group (N=16) |
|---|---|---|
| **SAEs** | | |
| SAEs — no. | 21 | 12 |
| Patients with ≥1 SAE — no. (%) | 10 (66.7) | 5 (31.3) |
| Blood and lymphatic system disorders — no. (%) | 4 (26.7) | 1 (6.3) |
| Anemia | 3 (20.0) | 1 (6.3) |
| Thrombocytopenia | 1 (6.7) | 0 |
| Cardiac disorders — no. (%) | 0 | 1 (6.3) |
| Cardio-respiratory arrest | 0 | 1 (6.3) |
| Cardiogenic shock | 0 | 1 (6.3) |
| Hepatobiliary disorders — no. (%) | 1 (6.7) | 0 |
| Cholecystitis | 1 (6.7) | 0 |
| Infections and infestations — no. (%) | 6 (40.0) | 3 (18.8) |
| Bacteremia | 1 (6.7) | 1 (6.3) |
| COVID-19 | 1 (6.7) | 0 |
| COVID-19 pneumonia | 2 (13.3) | 1 (6.3) |
| Pneumonia | 1 (6.7) | 0 |
| Pneumonia bacterial | 2 (13.3) | 0 |
| Septic shock | 0 | 2 (12.5) |
| Investigations — no. (%) | 1 (6.7) | 1 (6.3) |
| Liver function test increased | 1 (6.7) | 0 |
| Transaminases increased | 0 | 1 (6.3) |
| Metabolism and nutrition disorders — no. (%) | 1 (6.7) | 0 |
| Acidosis | 1 (6.7) | 0 |
| Hypokalemia | 1 (6.7) | 0 |
| Nervous system disorders — no. (%) | 2 (13.3) | 1 (6.3) |
| Alcoholic seizure | 0 | 1 (6.3) |
| Cerebrovascular accidence | 1 (6.7) | 0 |
| Seizure like phenomena | 1 (6.7) | 0 |
| Renal and urinary disorders — no. (%) | 2 (13.3) | 0 |
| Acute kidney injury | 2 (13.3) | 0 |
| Respiratory, thoracic, and mediastinal disorders — no. (%) | 2 (13.3) | 2 (12.5) |
| Acute respiratory distress syndrome | 0 | 1 (6.3) |
| Pneumothorax | 0 | 1 (6.3) |
| Respiratory acidosis | 1 (6.7) | 0 |
| Respiratory failure | 1 (6.7) | 0 |
| Vascular disorders | 0 | 1 (6.3) |
| Hypertension | 0 | 1 (6.3) |
| **AESIs** | | |
| AESIs — no. | 11 | 11 |
| Patients with ≥1 AESI — no. (%) | 7 (46.7) | 8 (50.0) |
| Blood and lymphatic system disorders — no. (%) | 0 | 1 (6.3) |
| Neutropenia | 0 | 1 (6.3) |
| Infections and infestations— no. (%) | 3 (20.0) | 4 (25.0) |
| Acinetobacter infection | 1 (6.7) | 0 |

| | | |
|---|---|---|
| Bacteremia | 2 (13.3) | 2 (12.5) |
| Pneumonia | 1 (6.7) | 1 (6.3) |
| Pneumonia bacterial | 1 (6.7) | 0 |
| Septic shock | 0 | 1 (6.3) |
| Staphylococcal infection | 0 | 1 (6.3) |
| Urinary tract infection | 1 (6.7) | 0 |
| Investigations — no. (%) | 3 (20.0) | 5 (31.3) |
| Alanine aminotransferase increased | 1 (6.7) | 0 |
| Aspartate aminotransferase increased | 1 (6.7) | 2 (12.5) |
| Hepatic enzyme increased | 0 | 1 (6.3) |
| Liver function test increased | 2 (13.3) | 1 (6.3) |
| Transaminases increased | 0 | 1 (6.3) |
| Nervous system disorders — no. (%) | 1 (6.7) | 0 |
| Seizure like phenomena | 1 (6.7) | 0 |

* AESI denotes adverse event of special interest, IV intravenous, SAE serious adverse event.

### Phase 3 (Cohort 3) Study Results

#### Phase 3 (Cohort 3): Flow Diagram

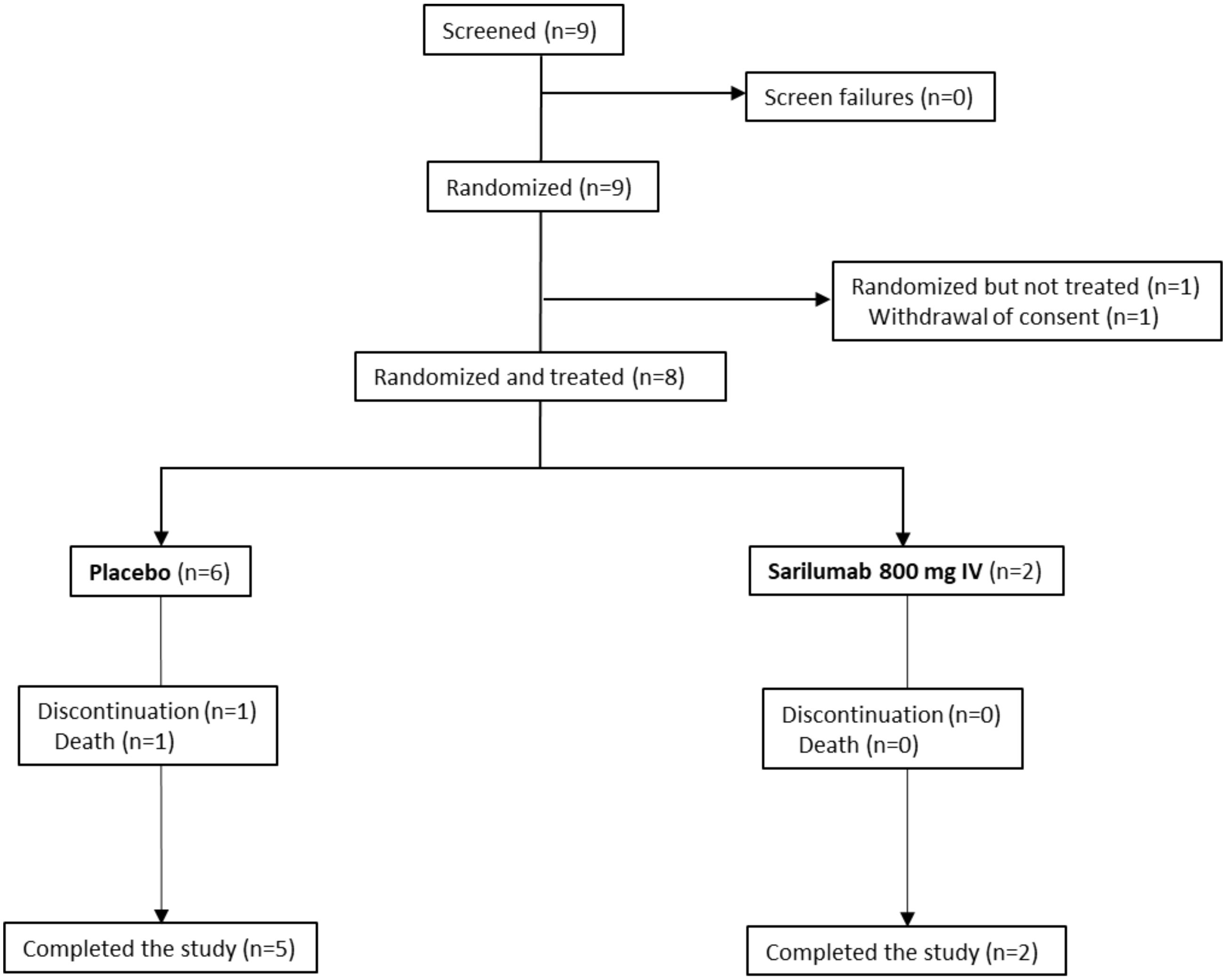

#### Phase 3 (Cohort 3): Demographics and Baseline Characteristics (ITT Population)*

| | Placebo Group (N=6) | Sarilumab 800 mg IV Group (N=2) |
|---|---|---|
| Median age — years (Q1:Q3) | 59.0 (55.0:69.0) | 66.0 (61.0:71.0) |
| Male sex — no. (%) | 3 (50.0) | 2 (100.0) |
| Ethnicity — no. (%) | | |
| Hispanic or Latino | 2 (33.3) | 1 (50.0) |
| Not Hispanic or Latino | 3 (50.0) | 0 |
| Not reported | 0 | 1 (50.0) |
| Unknown | 1 (16.7) | 0 |
| Race — no. (%) | | |
| White | 2 (33.3) | 1 (50.0) |
| Black or African American | 2 (33.3) | 0 |
| Asian | 0 | 0 |
| American Indian or Alaska Native | 0 | 0 |
| Native Hawaiian or Pacific Islander | 0 | 0 |
| Not reported | 2 (33.3) | 0 |
| Other | 0 | 1 (50.0) |
| Mean weight — kg | 99.02±13.53 | 91.60±4.53 |
| Body-mass index†, | 34.14±3.68 | 33.05±0.92 |
| CRP, mg/l, n‡, median (min:max) | 6, 162.500 (89.00:325.00) | 2, 258.081 (116.16:400.00) |
| IL-6, pg/ml, n‡, median (min:max) | 5, 45.590 (9.59:270.21) | 1, 124.260 (124.26:124.26) |
| Comorbidities — no. (%) | | |
| Hypertension | 4 (66.7) | 1 (50.0) |
| Diabetes | 1 (16.7) | 2 (100.0) |
| Obesity§ | 4 (66.7) | 2 (100.0) |
| Systemic corticosteroid use — no. (%) | 3 (50.0) | 0 |

*Plus–minus values are means ±SD. CRP denotes C-reactive protein, ITT intention-to-treat, IV intravenous, SD standard deviation.
†The body-mass index is the weight in kilograms divided by the square of the height in meters.
‡Patients with available data.
§BMI >30 $kg/m^2$.

#### Phase 3 (Cohort 3): Overview of Adverse Events (Safety Population)*

| | **Placebo Group (N=6)** | **Sarilumab 800 mg IV Group (N=2)** |
|---|---|---|
| TEAEs — total no. | 8 | 3 |
| SAEs — total no. | 7 | 1 |
| AESIs — total no. | 1 | 2 |
| Serious AESIs — total no. | 0 | 0 |
| Patients with any TEAE — no. (%) | 1 (16.7) | 2 (100.0) |
| Patients with any SAE — no. (%) | 1 (16.7) | 1 (50.0) |
| Patients with any AESI — no. (%) | 1 (16.7) | 2 (100.0) |
| Patients with any serious AESI — no. (%) | 0 | 0 |
| Patients with any TEAE leading to death — no. (%) | 1 (16.7) | 0 |
| Patients with any TEAE leading to withdrawal from the study drug — no. (%) | 0 | 0 |
| Patients with any TEAE leading to study infusion interruption — no. (%) | 0 | 1 (50.0) |
| Related TEAEs — no. | 1 | 1 |
| Related SAEs — no. | 0 | 0 |
| Related AESIs — no. | 1 | 1 |
| Related serious AESIs — no. | 0 | 0 |
| Patients with related TEAE — no. (%) | 1 (16.7) | 1 (50.0) |
| Patients with related SAE — no. (%) | 0 | 0 |
| Patients with related AESI — no. (%) | 1 (16.7) | 1 (50.0) |
| Patients with related serious AESI — no. (%) | 0 | 0 |

*AESI denotes adverse event of special interest, IV intravenous, TEAE treatment-emergent adverse event, SAE serious adverse event.

#### Phase 3 (Cohort 3): Treatment-Emergent Serious Adverse Events and Adverse Events of Special Interest Occurring in ≥5% of Patients in Any Group (Safety Population)*

| System Organ Class<br>Preferred Term | Placebo Group (N=6) | Sarilumab 800 mg IV Group (N=2) |
|---|---|---|
| **SAEs** | | |
| SAEs — no. | 7 | 1 |
| Patients with ≥1 SAE — no. (%) | 1 (16.7) | 1 (50.0) |
| Cardiac disorders — no. (%) | 1 (16.7) | 0 |
| Atrial fibrillation | 1 (16.7) | 0 |
| Pulseless electrical activity | 1 (16.7) | 0 |
| Infections and infestations — no. (%) | 1 (16.7) | 0 |
| Septic shock | 1 (16.7) | 0 |
| Renal and urinary disorders — no. (%) | 1 (16.7) | 0 |
| Acute kidney injury | 1 (16.7) | 0 |
| Respiratory, thoracic, and mediastinal disorders — no. (%) | 1 (16.7) | 1 (50.0) |
| Acute respiratory failure | 1 (16.7) | 1 (50.0) |
| Vascular disorders — no. (%) | 1 (16.7) | 0 |
| Deep vein thrombosis | 1 (16.7) | 0 |
| **AESIs** | | |
| AESIs — no. | 1 | 2 |
| Patients with ≥1 AESI — no. (%) | 1 (16.7) | 2 (100.0) |
| Infections and infestations — no. (%) | 1 (16.7) | 1 (50.0) |
| Staphylococcal bacteremia | 1 (16.7) | 0 |
| Staphylococcal infection | 0 | 1 (50.0) |
| Respiratory, thoracic, and mediastinal disorders — no. (%) | 0 | 1 (50.0) |
| Dyspnea | 0 | 1 (50.0) |

*AESI denotes adverse event of special interest, IV intravenous, SAE serious adverse event.

### Supplementary Figures

#### Figure S1. Schematic Overview of the Study Design (Phase 2 and Phase 3)

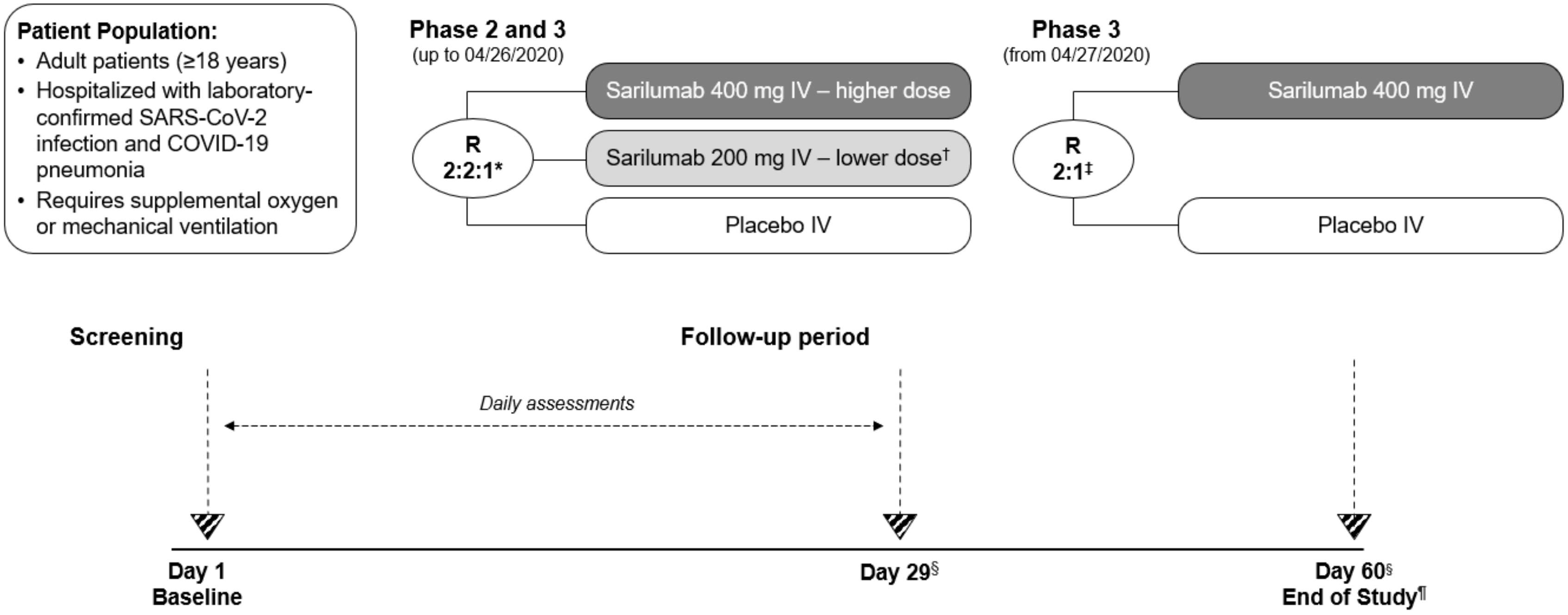

*Stratified by corticosteroid use and disease severity; †Following Phase 2 interim analysis, the IDMC recommended to eliminate the sarilumab 200 mg group and restrict future enrolment to critical patients; ‡Critical stratum only; §If the patient has been discharged from the hospital before day 29 or 60 the study site staff will contact the patient for a follow-up phone call; ¶The End of Study will be on day 60 or day of death, whichever comes first.

IDMC, Independent Data Monitoring Committee; IV, intravenous; R, randomized.

#### Figure S2. Phase 3 (Cohort 1): Flow Diagram

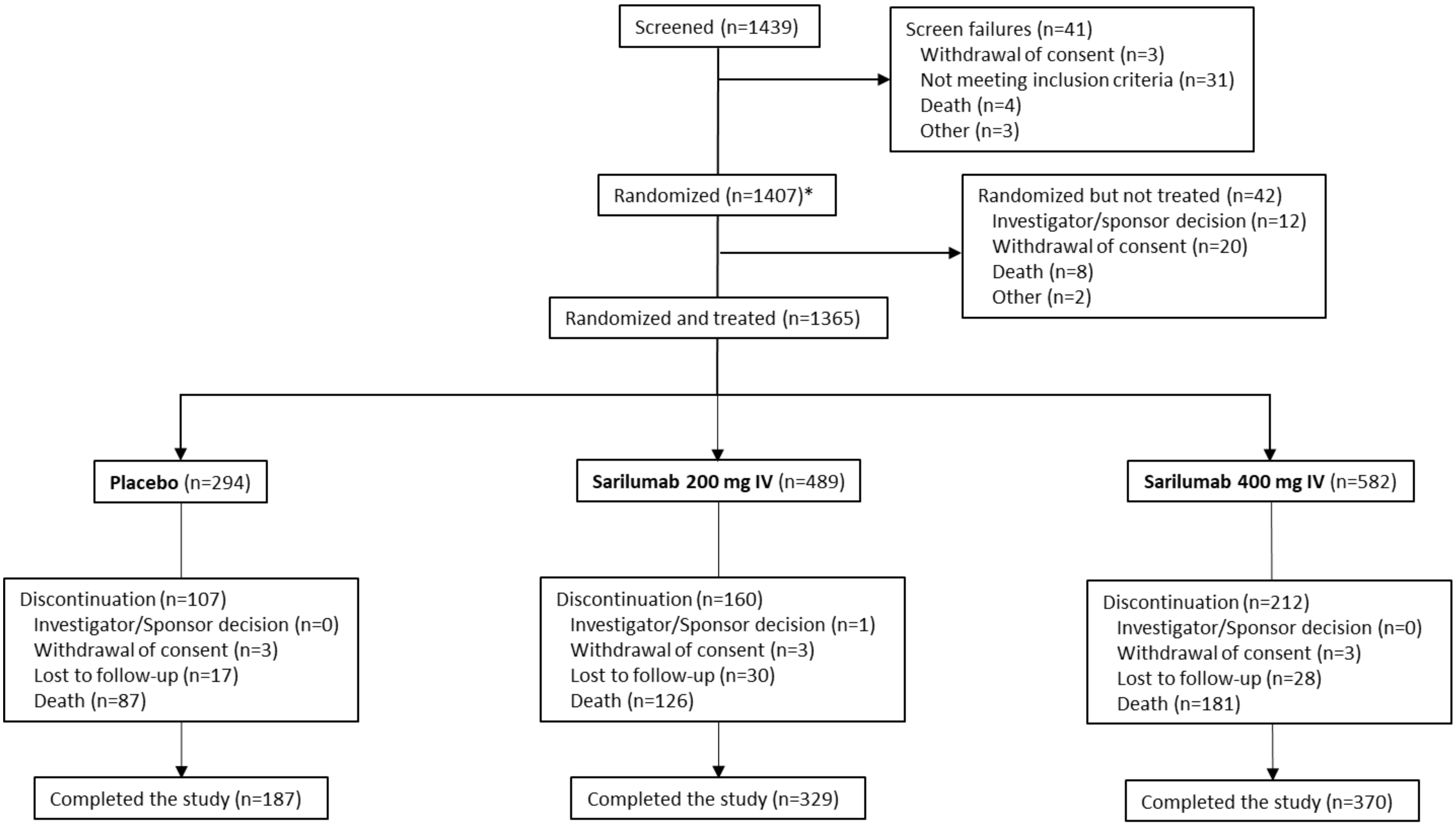

*Includes 9 screen failures that were randomized.

#### Figure S3. Phase 3 (Cohort 1): CRP Over Time in Critical Patients (Safety Population)

A. Critical patients receiving mechanical ventilation at baseline

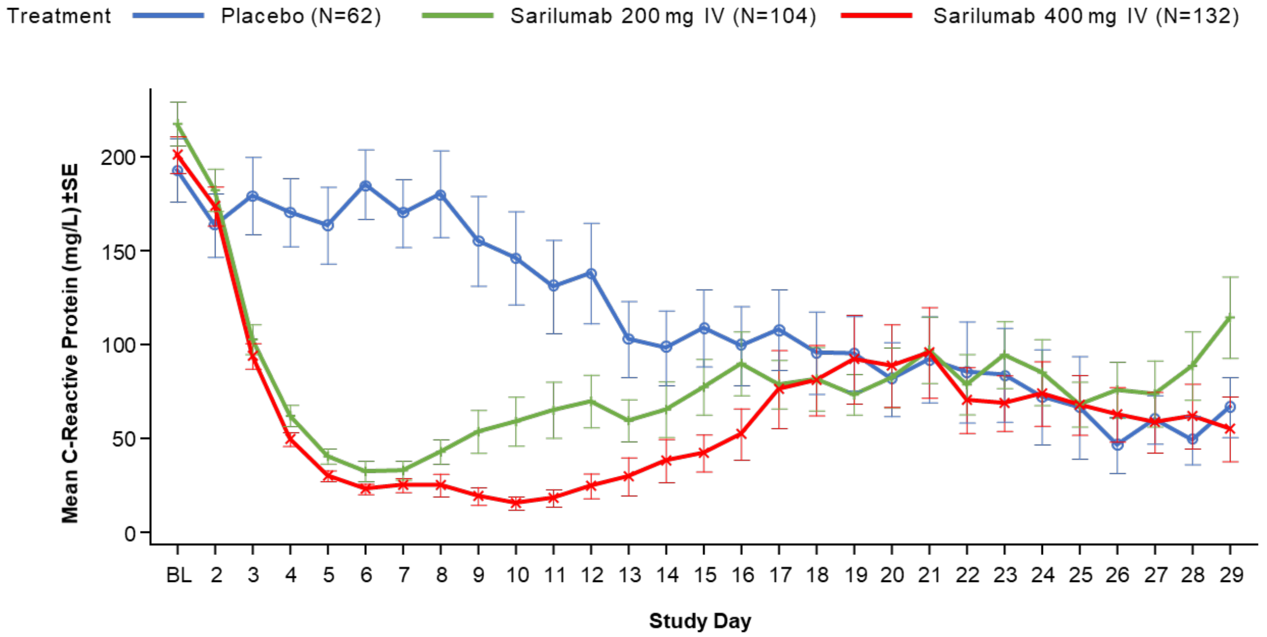

**Number of patients on each study day**

| | | | | | | | | | | | | | | | | | | | | | | | | | | | | | |
|---|---|---|---|---|---|---|---|---|---|---|---|---|---|---|---|---|---|---|---|---|---|---|---|---|---|---|---|---|---|
| Placebo (N=62) | 59 | 44 | 40 | 53 | 39 | 37 | 40 | 30 | 26 | 29 | 24 | 23 | 23 | 22 | 29 | 18 | 18 | 17 | 15 | 14 | 16 | 14 | 16 | 13 | 11 | 9 | 11 | 11 | 11 |
| Sarilumab 200 mg IV (N=104) | 100 | 81 | 73 | 90 | 73 | 62 | 74 | 59 | 51 | 49 | 39 | 43 | 39 | 32 | 38 | 31 | 32 | 29 | 27 | 26 | 22 | 25 | 22 | 23 | 20 | 17 | 20 | 21 | 18 |
| Sarilumab 400 mg IV (N=132) | 121 | 101 | 100 | 115 | 90 | 84 | 101 | 78 | 73 | 67 | 60 | 52 | 43 | 42 | 54 | 37 | 26 | 31 | 28 | 30 | 25 | 26 | 27 | 23 | 20 | 20 | 15 | 17 | 16 |

B. Critical patients **not** receiving mechanical ventilation at baseline

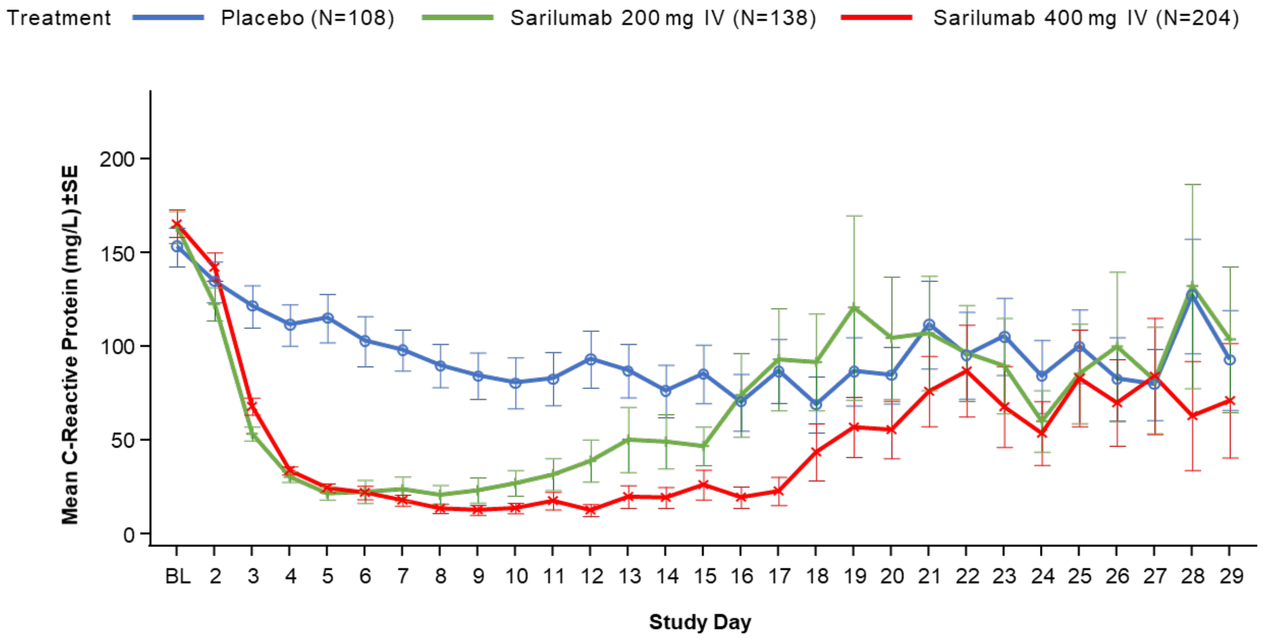

**Number of patients on each study day**

| | | | | | | | | | | | | | | | | | | | | | | | | | | | | | |
|---|---|---|---|---|---|---|---|---|---|---|---|---|---|---|---|---|---|---|---|---|---|---|---|---|---|---|---|---|---|
| Placebo (N=108) | 103 | 81 | 84 | 92 | 80 | 69 | 76 | 66 | 53 | 45 | 48 | 36 | 41 | 33 | 33 | 26 | 28 | 25 | 24 | 23 | 19 | 20 | 21 | 20 | 20 | 16 | 14 | 9 | 12 |
| Sarilumab 200 mg IV (N=138) | 123 | 101 | 87 | 109 | 83 | 71 | 81 | 57 | 52 | 42 | 38 | 40 | 28 | 28 | 33 | 16 | 19 | 19 | 12 | 16 | 15 | 15 | 17 | 13 | 9 | 6 | 8 | 7 | 10 |
| Sarilumab 400 mg IV (N=204) | 192 | 167 | 141 | 166 | 132 | 112 | 138 | 97 | 77 | 66 | 68 | 62 | 48 | 51 | 50 | 40 | 37 | 34 | 31 | 31 | 27 | 26 | 24 | 25 | 19 | 17 | 14 | 12 | 11 |

### Supplementary Tables

#### Table S1. Summary of Analysis Populations by Disease Severity Strata*

| | Placebo Group | Sarilumab 200 mg IV Group | Sarilumab 400 mg IV Group |
|---|---|---|---|
| **Phase 2** | | | |
| Intention-to-treat — no.† | | | |
| All disease severity | 90 | 187 | 180 |
| Severe‡ | 25 | 50 | 51 |
| Critical | 44 | 94 | 88 |
| MSOD/Immunocompromised‡ | 21 | 43 | 41 |
| Modified intention-to-treat — no. § | | | |
| All disease severity | 80 | 152 | 153 |
| Severe | 21 | 37 | 42 |
| Critical | 41 | 80 | 78 |
| MSOD/Immunocompromised | 18 | 35 | 33 |
| Safety population — no.¶ | | | |
| All disease severity | 90 | 187 | 180 |
| Severe | 25 | 50 | 51 |
| Critical | 44 | 94 | 88 |
| MSOD/Immunocompromised | 21 | 43 | 41 |
| **Phase 3 – Cohort 1** | | | |
| Intention-to-treat — no.† | | | |
| Severe | 70 | 140 | 137 |
| Critical | 170 | 242 | 338 |
| Critical receiving mechanical ventilation at baseline# | 62 | 104 | 132 |
| Critical not receiving mechanical ventilation at baseline | 108 | 138 | 204 |
| MSOD/Immunocompromised | 54 | 107 | 107 |
| Safety population — no.¶ | | | |
| Severe | 70 | 140 | 137 |
| Critical | 170 | 242 | 338 |
| Critical receiving mechanical ventilation at baseline# | 62 | 104 | 132 |
| Critical not receiving mechanical ventilation at baseline | 108 | 138 | 204 |
| MSOD/Immunocompromised | 54 | 107 | 107 |

| Integrated Phase (Phase 2 and Phase 3 [Cohort 1]) | | | |
|---|---|---|---|
| Intention-to-treat — no.† | | | |
| Severe | 95 | 190 | 188 |
| Critical | 214 | 335 | 423 |
| Critical receiving mechanical ventilation at baseline# | 80 | 164 | 179 |
| Critical <u>not</u> receiving mechanical ventilation at baseline | 134 | 171 | 244 |

*IV denotes intravenous, MSOD multiple system organ dysfunction.

†Intention-to-treat population includes all randomized patients who received at least one dose of the study drug.

‡Severe, MSOD, and immunocompromised populations were considered exploratory based on the statistical analysis plan.

§Modified intention-to-treat population includes randomized patients who received at least one dose of the study drug and had high baseline IL-6 levels.

¶Safety population includes all randomized patients who received at least one dose of the study drug.

#Pre-specified primary end point.

#### Table S2. Phase 3 (Cohort 1) – Severe and MSOD Strata: Demographics and Baseline Characteristics (EITT Population)*

| | Severe Patients | | | | MSOD Patients | | | |
|---|---|---|---|---|---|---|---|---|
| | **Placebo (N=70)** | **Sarilumab 200 mg IV Group (N=140)** | **Sarilumab 400 mg IV Group (N=137)** | **Total (N=347)** | **Placebo (N=46)** | **Sarilumab 200 mg IV Group (N=95)** | **Sarilumab 400 mg IV Group (N=92)** | **Total (N=233)** |
| Median age — years (Q1:Q3) | 63.5 (49.0:72.0) | 62.5 (50.0:72.0) | 63.0 (54.0:74.0) | 63.0 (51.0:73.0) | 60.5 (49.0:71.0) | 61.0 (53.0:67.0) | 64.5 (53.5:71.5) | 62.0 (53.0:69.0) |
| Male sex — no. (%) | 45 (64.3) | 81 (57.9) | 86 (62.8) | 212 (61.1) | 25 (54.3) | 54 (56.8) | 58 (63.0) | 137 (58.8) |
| Ethnicity— no. (%) | | | | | | | | |
| Hispanic or Latino | 19 (27.1) | 30 (21.4) | 30 (21.9) | 79 (22.8) | 12 (26.1) | 27 (28.4) | 21 (22.8) | 60 (25.8) |
| Not Hispanic or Latino | 40 (57.1) | 80 (57.1) | 88 (64.2) | 208 (59.9) | 26 (56.5) | 46 (48.4) | 47 (51.1) | 119 (51.1) |
| Not reported | 7 (10.0) | 20 (14.3) | 13 (9.5) | 40 (11.5) | 7 (15.2) | 16 (16.8) | 16 (17.4) | 39 (16.7) |
| Unknown | 4 (5.7) | 10 (7.1) | 6 (4.4) | 20 (5.8) | 1 (2.2) | 6 (6.3) | 8 (8.7) | 15 (6.4) |
| Race — no. (%) | | | | | | | | |
| White | 28 (40.0) | 45 (32.1) | 49 (35.8) | 122 (35.2) | 11 (23.9) | 29 (30.5) | 25 (27.2) | 65 (27.9) |
| Black or African American | 10 (14.3) | 35 (25.0) | 25 (18.2) | 70 (20.2) | 8 (17.4) | 23 (24.2) | 23 (25.0) | 54 (23.2) |
| Asian | 2 (2.9) | 6 (4.3) | 13 (9.5) | 21 (6.1) | 5 (10.9) | 3 (3.2) | 2 (2.2) | 10 (4.3) |
| American Indian or Alaska Native | 0 | 2 (1.4) | 0 | 2 (0.6) | 1 (2.2) | 0 | 0 | 1 (0.4) |
| Native Hawaiian or Pacific Islander | 1 (1.4) | 1 (0.7) | 1 (0.7) | 3 (0.9) | 0 | 0 | 1 (1.1) | 1 (0.4) |
| Not reported | 17 (24.3) | 36 (25.7) | 25 (18.2) | 78 (22.5) | 15 (32.6) | 34 (35.8) | 30 (32.6) | 79 (33.9) |
| Other | 12 (17.1) | 15 (10.7) | 24 (17.5 | 51 (14.7) | 6 (13.0) | 6 (6.3) | 11 (12.0) | 23 (9.9) |
| Mean weight — kg | 87.31±21.57 | 92.55±23.90 | 88.40±21.72 | 89.84±22.64 | 92.51±24.00 | 102.34±38.81 | 88.94±22.41 | 95.21±31.01 |
| Body-mass index†, | 31.83±8.85 | 32.59±7.80 | 30.68±6.90 | 31.68±7.72 | 32.54±8.50 | 35.20±12.64 | 31.14±7.32 | 33.13±10.22 |

| | | | | | | | | |
|---|---|---|---|---|---|---|---|---|
| CRP, mg/l, n‡, median (min:max) | 64, 96.201 (4.00:382.80) | 133, 105.502 (7.00:999.00) | 114, 103.400 (3.00:1260.00) | 311, 102.00 (3.00:1260.00) | 44, 179.004 (21.00:475.00) | 89, 195.404 (10.17:2626.00) | 86, 216.00 (16.00:723.00) | 219, 210.604 (10.17:2626.00) |
| IL-6, pg/ml, n‡, median (min:max) | 64, 53.570 (9.10:571.26) | 126, 52.585 (9.10:1713.25) | 117, 54.240 (9.10:696.18) | 307, 54.240 (9.10:1713.25) | 36, 179.765 (9.59:32374.39) | 74, 226.755 (9.59:17047.47) | 76, 306.430 (9.59:9488.14) | 186, 243.165 (9.59:32374.39) |
| Comorbidities — no. (%) | | | | | | | | |
| Hypertension | 37 (52.9) | 73 (52.1) | 74 (54.0) | 184 (53.0) | 26 (56.5) | 55 (57.9) | 45 (48.9) | 126 (54.1) |
| Diabetes | 14 (20.0) | 33 (23.6) | 26 (19.0) | 73 (21.0) | 8 (17.4) | 15 (15.8) | 17 (18.5) | 40 (17.2) |
| Obesity§ | 34 (48.6) | 75 (53.6) | 57 (41.6) | 166 (47.8) | 24 (52.2) | 60 (63.2) | 39 (42.2) | 123 (52.8) |
| Systemic corticosteroid use — no. (%) | 23 (32.9) | 47 (33.6) | 44 (32.1) | 114 (32.9) | 14 (30.4) | 30 (31.6) | 29 (31.5) | 73 (31.3) |

*Plus–minus values are means ±SD. CRP denotes C-reactive protein, EITT exploratory intention-to-treat, IL-6 interleukin-6, IV intravenous, SD standard deviation, Q quartile.

†The body-mass index is the weight in kilograms divided by the square of the height in meters.

‡Patients with available data.

§Body-mass index >30 $kg/m^2$.

### Table S3. Integrated Phase 2/3 Population: Demographics and Baseline Characteristics (ITT Population)*

| | Severe Patients | | | | Critical Patients Receiving Mechanical Ventilation at Baseline | | | | Critical Patients Not Receiving Mechanical Ventilation at Baseline | | | | All Critical Patients | | | |
|---|---|---|---|---|---|---|---|---|---|---|---|---|---|---|---|---|
| | Placebo Group (N=95) | Sarilumab 200 mg IV Group (N=190) | Sarilumab 400 mg IV Group (N=188) | Total (N=473) | Placebo Group (N=80) | Sarilumab 200 mg IV Group (N=164) | Sarilumab 400 mg IV Group (N=179) | Total (N=423) | Placebo Group (N=134) | Sarilumab 200 mg IV Group (N=171) | Sarilumab 400 mg IV Group (N=244) | Total (N=549) | Placebo Group (N=214) | Sarilumab 200 mg IV Group (N=335) | Sarilumab 400 mg IV Group (N=423) | Total (N=972) |
| Median age — years (Q1:Q3) | 63.0 (50.0:71.0) | 61.0 (48.0:71.0) | 61.0 (50.0:72.0) | 61.0 (50.0:72.0) | 58.5 (48.5:69.0) | 59.0 (49.5:67.0) | 60.0 (50.0:70.0) | 59.0 (49.0:69.0) | 60.5 (52.0:71.0) | 59.0 (50.0:69.0) | 62.5 (51.0:71.0) | 61.0 (51.0:70.0) | 60.0 (50.0:69.0) | 59.0 (50.0:68.0) | 62.0 (51.0:71.0) | 60.0 (50.0:69.0) |
| Male sex — no. (%) | 60 (63.2) | 115 (60.5) | 124 (66.0) | 299 (63.2) | 59 (73.8) | 109 (66.5) | 123 (68.7) | 291 (68.8) | 89 (66.4) | 127 (74.3) | 173 (70.9) | 389 (70.9) | 148 (69.2) | 236 (70.4) | 296 (70.0) | 680 (70.0) |
| Ethnicity — no. (%) | | | | | | | | | | | | | | | | |
| Hispanic or Latino | 23 (24.2) | 43 (22.6) | 39 (20.7) | 105 (22.2) | 28 (35.0) | 51 (31.1) | 52 (29.1) | 131 (31.0) | 38 (28.4) | 55 (32.2) | 83 (34.0) | 176 (32.1) | 66 (30.8) | 106 (31.6) | 135 (31.9) | 307 (31.6) |
| Not Hispanic or Latino | 56 (58.9) | 109 (57.4) | 122 (64.9) | 287 (60.7) | 37 (46.3) | 79 (48.2) | 98 (54.7) | 214 (50.6) | 80 (59.7) | 83 (48.5) | 111 (45.5) | 274 (49.9) | 117 (54.7) | 162 (48.4) | 209 (49.4) | 488 (50.2) |
| Not reported | 11 (11.6) | 23 (12.1) | 20 (10.6) | 54 (11.4) | 6 (7.5) | 14 (8.5) | 17 (9.5) | 37 (8.7) | 11 (8.2) | 21 (12.3) | 29 (11.9) | 61 (11.1) | 17 (7.9) | 35 (10.4) | 46 (10.9) | 98 (10.1) |
| Unknown | 5 (5.3) | 15 (7.9) | 7 (3.7) | 27 (5.7) | 9 (11.3) | 20 (12.2) | 12 (6.7) | 41 (9.7) | 5 (3.7) | 12 (7.0) | 21 (8.6) | 38 (6.9) | 14 (6.5) | 32 (9.6) | 33 (7.8) | 79 (8.1) |
| Race — no. (%) | | | | | | | | | | | | | | | | |
| White | 43 (45.3) | 67 (35.3) | 71 (37.8) | 181 (38.3) | 38 (47.5) | 66 (40.2) | 71 (39.7) | 175 (41.4) | 52 (38.8) | 60 (35.1) | 84 (34.4) | 196 (35.7) | 90 (42.1) | 126 (37.6) | 155 (36.6) | 371 (38.2) |
| Black or African American | 14 (14.7) | 43 (22.6) | 33 (17.6) | 90 (19.0) | 7 (8.8) | 26 (15.9) | 38 (21.2) | 71 (16.8) | 22 (16.4) | 26 (15.2) | 41 (16.8) | 89 (16.2) | 29 (13.6) | 52 (15.5) | 79 (18.7) | 160 (16.5) |
| Asian | 3 (3.2) | 13 (6.8) | 19 (10.1) | 35 (7.4) | 8 (10.0) | 11 (6.7) | 10 (5.6) | 29 (6.9) | 11 (8.2) | 8 (4.7) | 8 (3.3) | 27 (4.9) | 19 (8.9) | 19 (5.7) | 18 (4.3) | 56 (5.8) |
| American Indian or Alaska Native | 0 | 2 (1.1) | 0 | 2 (0.4) | 0 | 1 (0.6) | 0 | 1 (0.2) | 1 (0.7) | 0 | 1 (0.4) | 2 (0.4) | 1 (0.5) | 1 (0.3) | 1 (0.2) | 3 (0.3) |
| Native Hawaiian or Pacific Islander | 1 (1.1) | 1 (0.5) | 2 (1.1) | 4 (0.8) | 0 | 0 | 0 | 0 | 0 | 1 (0.6) | 0 | 1 (0.2) | 0 | 1 (0.3) | 0 | 1 (0.1) |
| Not reported | 20 (21.1) | 44 (23.2) | 32 (17.0) | 96 (20.3) | 20 (25.0) | 44 (26.8) | 42 (23.5) | 106 (25.1) | 24 (17.9) | 46 (26.9) | 76 (31.1) | 146 (26.6) | 44 (20.6) | 90 (26.9) | 118 (27.9) | 252 (25.9) |
| Other | 14 (14.7) | 20 (10.5) | 31 (16.5) | 65 (13.7) | 7 (8.8) | 16 (9.8) | 18 (10.1) | 41 (9.7) | 24 (17.9) | 30 (17.5) | 34 (13.9) | 88 (16.0) | 31 (14.5) | 46 (13.7) | 52 (12.3) | 129 (13.3) |
| Mean weight — kg | 87.13±21.34 | 92.22±23.39 | 88.83±22.47 | 89.83±22.66 | 87.43±20.27 | 90.26±21.85 | 95.25±27.61 | 91.90±24.40 | 90.48±22.75 | 89.13±22.60 | 88.81±23.23 | 89.32±22.89 | 89.35±21.86 | 89.69±22.21 | 91.61±25.4) | 90.46±23.59 |
| Body-mass index† | 31.33±8.23 | 32.26±7.80 | 30.65±7.16 | 31.44±7.67 | 30.58±6.70 | 31.68±7.21 | 32.73±8.66 | 31.93±7.82 | 31.48±7.57 | 30.85±7.60 | 31.33±7.06 | 31.22±7.34 | 31.14±7.25 | 31.26±7.41 | 31.94±7.82 | 31.53±7.56 |
| CRP, mg/l, n‡, median (min:max) | 89, 102.000 (4.00:382.80) | 181, 105.480 (7.00:999.00) | 163, 108.202 (1.00:1260.00) | 433, 105.000 (1.00:1260.00) | 77, 193.780 (9.00:603.01) | 158, 198.320 (5.00:591.00) | 167, 185.700 (8.68:662.00) | 402, 193.890 (5.00:662.00) | 127, 141.503 (1.70:476.81) | 156, 157.102 (5.00:819.94) | 230, 163.900 (1.20:454.21) | 513, 152.000 (1.20:819.94) | 204, 152.500 (1.70:603.01) | 314, 180.122 (5.00:819.94) | 397, 173.803 (1.20:662.00) | 915, 173.003 (1.20:819:94) |
| IL-6, pg/ml, n‡, median (min:max) | 86, 61.585 (9.10:571.26) | 169, 53.350 (9.10:1713.25) | 162, 58.250 (9.10:2771.49) | 417, 58.360 (9.10:2771.49) | 73, 163.860 (9.10:2324.82) | 143, 169.540 (9.59:6531.58) | 156, 164.385 (9.59:21545.37) | 372, 164.695 (9.10:21545.37) | 127, 67.760 (9.59:2176.98) | 150, 67.795 (6.82:1436.60) | 213, 95.210 (9.10:1032.44) | 490, 77.790 (6.82:2176.98) | 200, 85.610 (9.10:2324.82) | 293, 116.140 (6.82:6531.58) | 369, 125.990 (9.10:21545.37) | 862, 114.375 (6.82:21545.37) |
| Comorbidities — no. (%) | | | | | | | | | | | | | | | | |
| Hypertension | 46 (48.4) | 93 (48.9) | 100 (53.2) | 239 (50.5) | 36 (45.0) | 69 (42.1) | 88 (49.2) | 193 (45.6) | 80 (59.7) | 82 (48.0) | 132 (54.1) | 294 (53.6) | 116 (54.2) | 151 (45.1) | 220 (52.0) | 487 (50.1) |
| Diabetes | 19 (20.0) | 38 (20.0) | 33 (17.6) | 90 (19.0) | 12 (15.0) | 38 (23.2) | 27 (15.1) | 77 (18.2) | 32 (23.9) | 36 (21.1) | 41 (16.8) | 109 (19.9) | 44 (20.6) | 74 (22.1) | 68 (16.1) | 186 (19.1) |
| Obesity§ | 43 (45.3) | 98 (51.6) | 75 (39.9) | 216 (45.7) | 36 (45.0) | 71 (43.3) | 96 (53.6) | 203 (48.0) | 61 (45.5) | 67 (39.2) | 116 (47.5) | 244 (44.4) | 97 (45.3) | 138 (41.2) | 212 (50.1) | 447 (46.0) |
| Systemic corticosteroid use — no. (%) | 26 (27.4) | 53 (27.9) | 50 (26.6) | 129 (27.3) | 22 (27.5) | 37 (22.6) | 50 (27.9) | 109 (25.8) | 47 (35.1) | 57 (33.3) | 87 (35.7) | 191 (34.8) | 69 (32.2) | 94 (28.1) | 137 (32.4) | 300 (30.9) |

*Plus–minus values are means ±SD. CRP denotes C-reactive protein, EITT exploratory intention-to-treat, IL-6 interleukin-6, IV intravenous, SD standard deviation, Q quartile.

†The body-mass index is the weight in kilograms divided by the square of the height in meters.

‡Patients with available data.

§BMI >30 kg/m².

#### Table S4. Phase 3 (Cohort 1): Summary of Primary and Selected Secondary Efficacy End Points in Critical Strata*

| Outcome | Critical Patients Receiving Mechanical Ventilation at Baseline (ITT) | | | | | Critical Patients Not Receiving Mechanical Ventilation at Baseline (ITT) | | | | | All Critical Patients (ITT) | | | | |
|---|---|---|---|---|---|---|---|---|---|---|---|---|---|---|---|
| | Placebo Group (N=62) n (%) [95% CI] | Sarilumab 200 mg IV Group (N=104) n (%) [95% CI] | Sarilumab 400 mg IV Group (N=132) n (%) [95% CI] | Risk difference Sarilumab 200 mg IV vs placebo [95% CI] | Risk difference Sarilumab 400 mg IV vs placebo [95% CI] | Placebo Group (N=108) n (%) [95% CI] | Sarilumab 200 mg IV Group (N=138) n (%) [95% CI] | Sarilumab 400 mg IV Group (N=204) n (%) [95% CI] | Risk difference Sarilumab 200 mg IV vs placebo [95% CI] | Risk difference Sarilumab 400 mg IV vs placebo [95% CI] | Placebo Group (N=170) n (%) [95% CI] | Sarilumab 200 mg IV Group (N=242) n (%) [95% CI] | Sarilumab 400 Group mg IV (N=338) n (%) [95% CI] | Risk difference Sarilumab 200 mg IV vs placebo [95% CI] | Risk difference Sarilumab 400 mg IV vs placebo [95% CI] |
| Proportion with 1-point improvement (day 22) | 22 (35.5%) [23.6, 47.4] | 45 (43.3%) [33.7, 52.8] | 57 (43.2%) [34.7, 51.6] | +7.1% [-8.4, 21.7] | +7.5% [-7.4, 21.3][†] | 70 (64.8%) [55.8, 73.8] | 92 (66.7%) [58.8, 74.5] | 117 (57.4%) [50.6, 64.1] | +2.0% [-9.7, 13.9] | -7.5% [-18.3, 3.9] | 92 (54.1%) [46.6, 61.6] | 137 (56.6%) [50.4, 62.9] | 174 (51.5%) [46.2, 56.8] | +2.7% [-7.0, 12.4] | -2.6% [-11.7, 6.5] |
| Proportion who died (day 29) | 26 (41.9%) [29.7, 54.2] | 35 (33.7%) [24.6, 42.7] | 48 (36.4%) [28.2, 44.6] | -7.7% [-22.8, 7.2] | -5.5% [-20.2, 8.7] | 17 (15.7%) [8.9, 22.6] | 25 (18.1%) [11.7, 24.5] | 54 (26.5%) [20.4, 32.5] | +2.3% [-7.5, 11.5] | +10.7% [0.9, 19.3] | 43 (25.3%) [18.8, 31.8] | 60 (24.8%) [19.4, 30.2] | 103 (30.5%) [25.6, 35.4] | -0.6% [-9.3, 7.7] | +5.2% [-3.3, 13.0] |
| Proportion who died (day 60) | 32 (51.6%) [39.2, 64.1] | 39 (37.5%) [28.2, 46.8] | 52 (39.4%) [31.1, 47.7] | -13.3% [-28.2, 2.3] | -11.9% [-26.4, 2.9] | 27 (25.0%) [16.8, 33.2] | 31 (22.5%) [15.5, 29.4] | 61 (29.9%) [23.6, 36.2] | -2.6% [-13.5, 7.9] | +4.8 [-5.8, 14.6] | 59 (34.7%) [27.6, 41.9] | 70 (28.9%) [23.2, 34.6] | 114 (33.7%) [28.7, 38.8] | -5.8% [-15.0, 3.2] | -1.0% [-9.8, 7.6] |
| Proportion who | 16 (25.8%) | 32 (30.8%) | 42 (31.8%) | +5.2% [-9.4, 18.6] | +5.7% [-8.4, 18.2] | 69 (63.9%) | 88 (63.8%) | 111 (54.4%) | +0.1% [-11.7, 12.2] | -9.5% [-20.4, 2.0] | 85 (50.0%) | 120 (49.6%) | 153 (45.3%) | +0.2% [-9.5, 9.9] | -4.7% [-13.8, 4.4] |

| | | | | | | | | | | | | | | | |
|---|---|---|---|---|---|---|---|---|---|---|---|---|---|---|---|
| recover (day 22)* | [14.9, 36.7] | [21.9, 39.6] | [23.9, 39.8] | | | [54.8, 72.9] | [55.7, 71.8] | [47.6, 61.2] | | | [42.5, 57.5] | [43.3, 55.9] | [40.0, 50.6] | | |
| Proportion alive not requiring MV/ECMO (day 22) | 22 (35.5 %) [23.6, 47.4] | 48 (46.2 %) [36.6, 55.7] | 59 (44.7 %) [36.2, 53.2] | +9.8% [-5.8, 24.3] | +8.8% [-6.1, 22.6] | 78 (72.2 %) [63.8, 80.7] | 106 (76.8 %) [69.8, 83.9] | 130 (63.7 %) [57.1, 70.3] | +4.8% [-6.0, 15.9] | -8.6% [-18.8, 2.4] | 100 (58.8 %) [51.4, 66.2] | 154 (63.6 %) [57.6, 69.7] | 189 (55.9 %) [50.6, 61.2] | +5.2% [-4.3, 14.7] | -2.9% [-11.8, 6.2] |
| Proportion with 2-point improveme nt (day 22) | 21 (33.9 %) [22.1, 45.7] | 40 (38.5 %) [29.1, 47.8] | 53 (40.2 %) [31.8, 48.5] | +4.1% [-11.2, 18.5] | +5.9 [-8.9, 19.5] | 68 (63.0 %) [53.9, 72.1] | 87 (63.0 %) [55.0, 71.1] | 108 (52.9 %) [46.1, 59.8] | +0.3% [-11.6, 12.4] | - 10.1% [-21.1, 1.4] | 89 (52.4 %) [44.8, 59.9] | 127 (52.5 %) [46.2, 58.8] | 161 (47.6 %) [42.3, 53.0] | +0.5% [-9.2, 10.2] | -4.7% [-13.8, 4.5] |
| Proportion discharged alive (day 22) | 14 (22.6 %) [12.2, 33.0] | 28 (26.9 %) [18.4, 35.4] | 35 (26.5 %) [19.0, 34.0] | +4.5% [-9.7, 17.3] | +3.7% [-10.0, 5.6] | 67 (62.0 %) [52.9, 71.2] | 87 (63.0 %) [55.0, 71.1] | 109 (53.4 %) [46.6, 60.3] | +1.2% [-10.8, 13.3] | -8.6% [-19.6, 3.0] | 81 (47.6 %) [40.1, 55.2] | 115 (47.5 %) [41.2, 53.8] | 144 (42.6 %) [37.3, 47.9] | +0.4% [-9.3, 10.1] | -5.0% [-14.1, 4.1] |

*ECMO denotes extracorporeal membrane oxygenation, IV intravenous, MV mechanical ventilation.
*Discharged, or alive without supplemental oxygen us or at pre-COVID-19 oxygen use.
†P value = 0.3261
Green = primary end point; blue = key secondary end point; orange = other secondary end points.
Positive risk differences indicate an effect favoring sarilumab for all end points except for the proportion who die by day 29 or day 60.

#### Table S5. Phase 3 (Cohort 1): Summary of Primary and Selected Secondary Efficacy End Points in Severe and MSOD Strata*

| | Severe patients (EITT) | | | | | MSOD patients (EITT) | | | | |
|---|---|---|---|---|---|---|---|---|---|---|
| Outcome | Placebo Group (N=70) no. (%) [95% CI] | Sarilumab 200 mg IV Group (N=140) no. (%) [95% CI] | Sarilumab 400 mg IV Group (N=137) no. (%) [95% CI] | Risk difference Sarilumab 200 mg IV vs placebo [95% CI] | Risk difference Sarilumab 400 mg IV vs placebo [95% CI] | Placebo Group (N=46) no. (%) [95% CI] | Sarilumab 200 mg IV Group (N=95) no. (%) [95% CI] | Sarilumab 400 mg IV Group (N=92) no. (%) [95% CI] | Risk difference Sarilumab 200 mg IV vs placebo [95% CI] | Risk difference Sarilumab 400 mg IV vs placebo [95% CI] |
| Proportion with 1-point improvement (day 22) | 58 (82.9%) [74.0–91.7] | 116 (82.9%) [76.6–89.1] | 108 (78.8%) [72.0–85.7] | -0.0% [-10.1–11.7] | -4.0% [-14.4–8.0] | 16 (34.8%) [21.0–48.5] | 36 (37.9%) [28.1–47.7] | 30 (32.6%) [23.0–42.2] | +3.1% [-14.0–18.8] | -2.2% [-19.1–13.6] |
| Proportion who died (day 29) | 8 (11.4%) [4.0–18.9] | 11 (7.9%) [3.4–12.3] | 16 (11.7%) [6.3–17.1] | -3.6% [-13.7–4.4] | +0.3% [-10.2–8.8] | 13 (28.3%) [15.2–41.3] | 36 (37.9%) [28.1–47.7] | 38 (41.3%) [31.2–51.4] | +9.6% [-7.3–24.5] | +13.0% [-4.2–28.0] |
| Proportion who died (day 60) | 9 (12.9%) [5.0–20.7] | 12 (8.6%) [3.9–13.2] | 21 (15.3%) [9.3–21.4] | -4.3% [-14.7–4.1] | +2.6% [-8.4–11.7] | 16 (34.8%) [21.0–48.5] | 40 (42.1%) [32.2–52.0] | 40 (43.5%) [33.3–53.6] | +7.3% [-10.0–23.0] | +8.6% [-8.7–24.5] |
| Proportion who recover (day 22)[†] | 59 (84.3%) [75.8–92.8] | 117 (83.6%) [77.4–89.7] | 109 (79.6%) [72.8–86.3] | -0.8% [-10.5–10.8] | -4.7% [-14.8–7.1] | 11 (23.9%) [11.6–36.2] | 29 (30.5%) [21.3–39.8] | 22 (23.9%) [15.2–32.6] | +6.6% [-9.7–20.8] | -0.1% [-16.1–13.8] |
| Proportion alive not requiring MV/ECMO (day 22) | 60 (85.7%) [77.5–93.9] | 126 (90.0%) [85.0–95.0] | 115 (83.9%) [77.8, 90.1] | +4.2% [-4.6–15.1] | -1.8% [-11.3–9.5] | 17 (37.0%) [23.0–50.9] | 38 (40.0%) [30.1–49.9] | 32 (34.8%) [25.1, 44.5] | +3.0% [-14.2–19.0] | -2.3% [-19.3–13.8] |

| | | | | | | | | | | |
|---|---|---|---|---|---|---|---|---|---|---|
| Proportion with 2-point improvement (day 22) | 57 (81.4%) [72.3–90.5] | 114 (81.4%) [75.0–87.9] | 108 (78.8%) [72.0–85.7] | -0.1% [-10.4–12.0] | -2.6% [-13.2–9.6] | 14 (30.4%) [17.1–43.7] | 33 (34.7%) [25.2–44.3] | 28 (30.4%) [21.0, 39.8] | +4.3% [-12.6–19.5] | -0.0% [-16.7–15.2] |
| Proportion discharged alive (day 22) | 58 (82.9%) [74.0–91.7] | 115 (82.1%) [75.8–88.5] | 108 (78.8%) [72.0–85.7] | -0.8% [-10.8–11.1] | -4.0% [-14.4–8.0] | 11 (23.9%) [11.6–36.2] | 24 (25.3%) [16.5–34.0] | 16 (17.4%) [9.6–25.1] | +1.3% [-14.6–15.3] | -6.7% [-22.1–6.8] |

*Positive risk differences indicate an effect favoring sarilumab for all end points except for the proportion who die by day 29 or day 60. ECMO denotes extracorporeal membrane oxygenation, IV intravenous, MV mechanical ventilation.

†Discharged, or alive without supplemental oxygen us or at pre–COVID-19 oxygen use.

#### Table S6. Phase 3 (Cohort 1) – Critical Stratum: Overview of Adverse Events (Safety Population)*

| | Critical patients receiving mechanical ventilation at baseline | | | Critical patients not receiving mechanical ventilation at baseline | | | All critical patients | | |
|---|---|---|---|---|---|---|---|---|---|
| | Placebo Group (N=62) | Sarilumab 200 mg IV Group (N=104) | Sarilumab 400 mg IV Group (N=132) | Placebo Group (N=108) | Sarilumab 200 mg IV Group (N=138) | Sarilumab 400 mg IV Group (N=204) | Placebo Group (N=170) | Sarilumab 200 mg IV Group (N=242) | Sarilumab 400 mg IV Group (N=338) |
| TEAEs — total no. | 139 | 217 | 259 | 130 | 176 | 280 | 269 | 393 | 544 |
| SAEs — total no. | 103 | 143 | 174 | 96 | 114 | 183 | 199 | 257 | 359 |
| AESIs — total no. | 61 | 102 | 128 | 49 | 89 | 124 | 110 | 191 | 255 |
| Serious AESIs — total no. | 25 | 28 | 43 | 15 | 27 | 27 | 40 | 55 | 70 |
| Patients with any TEAE — no. (%) | 51 (82.3) | 81 (77.9) | 106 (80.3) | 57 (52.8) | 76 (55.1) | 125 (61.3) | 108 (63.5) | 157 (64.9) | 233 (68.9) |
| Patients with any SAE — no. (%) | 44 (71.0) | 65 (62.5) | 88 (66.7) | 45 (41.7) | 54 (39.1) | 93 (45.6) | 89 (52.4) | 119 (49.2) | 183 (54.1) |
| Patients with any AESI — no. (%) | 31 (50.0) | 56 (53.8) | 71 (53.8) | 30 (27.8) | 51 (37.0) | 77 (37.7) | 61 (35.9) | 107 (44.2) | 150 (44.4) |
| Patients with any serious AESI — no. (%) | 14 (22.6) | 21 (20.2) | 32 (24.2) | 10 (9.3) | 16 (11.6) | 19 (9.3) | 24 (14.1) | 37 (15.3) | 51 (15.1) |
| Patients with any TEAE leading to death — no. (%) | 32 (51.6) | 38 (36.5) | 50 (37.9) | 27 (25.0) | 31 (22.5) | 59 (28.9) | 59 (34.7) | 69 (28.5) | 110 (32.5) |
| Patients with any TEAE leading to withdrawal from the study drug — no. (%) | 2 (3.2) | 5 (4.8) | 4 (3.0) | 4 (3.7) | 0 | 3 (1.5) | 6 (3.5) | 5 (2.1) | 7 (2.1) |
| Patients with any TEAE leading to study infusion interruption — no. (%) | 5 (8.1) | 5 (4.8) | 13 (9.8) | 2 (1.9) | 3 (2.2) | 5 (2.5) | 7 (4.1) | 8 (3.3) | 18 (5.3) |
| Related TEAEs — total no. | 25 | 57 | 77 | 36 | 42 | 71 | 61 | 99 | 150 |
| Related SAEs — total no. | 8 | 21 | 24 | 14 | 10 | 28 | 22 | 31 | 53 |
| Related AESIs — total no. | 24 | 43 | 69 | 27 | 41 | 63 | 51 | 84 | 133 |
| Related serious AESIs — total no. | 7 | 7 | 16 | 5 | 9 | 20 | 12 | 16 | 36 |
| Patients with related TEAE — no. (%) | 17 (27.4) | 31 (29.8) | 46 (34.8) | 21 (19.4) | 26 (18.8) | 48 (23.5) | 38 (22.4) | 57 (23.6) | 96 (28.4) |
| Patients with related SAE — no. (%) | 8 (12.9) | 13 (12.5) | 18 (13.6) | 9 (8.3) | 7 (5.1) | 19 (9.3) | 17 (10.0) | 20 (8.3) | 38 (11.2) |

| | | | | | | | | | |
|---|---|---|---|---|---|---|---|---|---|
| Patients with related AESI — no. (%) | 16 (25.8) | 28 (26.9) | 44 (33.3) | 17 (15.7) | 25 (18.1) | 45 (22.1) | 33 (19.4) | 53 (21.9) | 90 (26.6) |
| Patients with related serious AESI — no. (%) | 7 (11.3) | 6 (5.8) | 15 (11.4) | 3 (2.8) | 6 (4.3) | 14 (6.9) | 10 (5.9) | 12 (5.0) | 29 (8.6) |

*AESI denotes adverse event of special interest, IV intravenous, TEAE treatment-emergent adverse event, SAE serious adverse event.

**Table S7. Phase 3 (Cohort 1) – Severe and MSOD Strata: Overview of Adverse Events (Safety Population)***

| | Severe Patients | | | MSOD Patients | | |
|---|---|---|---|---|---|---|
| | Placebo Group (N=70) | Sarilumab 200 mg IV Group (N=140) | Sarilumab 400 mg IV Group (N=137) | Placebo Group (N=46) | Sarilumab 200 mg IV Group (N=95) | Sarilumab 400 mg IV Group (N=92) |
| TEAEs — total no. | 28 | 58 | 100 | 86 | 187 | 181 |
| SAEs — total no. | 22 | 38 | 71 | 39 | 125 | 117 |
| AESIs — total no. | 8 | 24 | 45 | 54 | 81 | 90 |
| Serious AESIs — total no. | 2 | 4 | 16 | 7 | 19 | 26 |
| Patients with any TEAE — no. (%) | 14 (20.0) | 36 (25.7) | 47 (34.3) | 37 (80.4) | 74 (77.9) | 73 (79.3) |
| Patients with any SAE — no. (%) | 11 (15.7) | 23 (16.4) | 30 (21.9) | 27 (58.7) | 67 (70.5) | 58 (63.0) |
| Patients with any AESI — no. (%) | 5 (7.1) | 20 (14.3) | 31 (22.6) | 28 (60.9) | 45 (47.4) | 51 (55.4) |
| Patients with any serious AESI — no.(%) | 1 (1.4) | 3 (2.1) | 10 (7.3) | 7 (15.2) | 14 (14.7) | 21 (22.8) |
| Patients with any TEAE leading to death — no. (%) | 9 (12.9) | 12 (8.6) | 19 (13.9) | 16 (34.8) | 40 (42.1) | 39 (42.4) |
| Patients with any TEAE leading to withdrawal from the study drug — no. (%) | 1 (1.4) | 0 | 0 | 3 (6.5) | 2 (2.1) | 1 (1.1) |
| Patients with any TEAE leading to study infusion interruption — no. (%) | 0 | 0 | 1 (0.7) | 0 | 1 (1.1) | 3 (3.3) |
| Related TEAEs — total no. | 2 | 23 | 18 | 30 | 36 | 40 |
| Related SAEs — total no. | 0 | 9 | 6 | 2 | 11 | 14 |
| Related AESIs — total no. | 2 | 14 | 16 | 30 | 33 | 36 |
| Related serious AESIs — total no. | 0 | 0 | 4 | 2 | 8 | 10 |
| Patients with related TEAE — no. (%) | 2 (2.9) | 13 (9.3) | 15 (10.9) | 17 (37.0) | 23 (24.2) | 24 (26.1) |
| Patients with related SAE — no. (%) | 0 | 3 (2.1) | 5 (3.6) | 2 (4.3) | 7 (7.4) | 10 (10.9) |
| Patients with related AESI — no. (%) | 2 (2.9) | 11 (7.9) | 14 (10.2) | 17 (37.0) | 23 (24.2) | 23 (25.0) |
| Patients with related serious AESI — no. (%) | 0 | 0 | 3 (2.2) | 2 (4.3) | 6 (6.3) | 9 (9.8) |

*AESI denotes adverse event of special interest, IV intravenous, TEAE treatment-emergent adverse event, SAE serious adverse event.

#### Table S8. Phase 3 (Cohort 1) – Severe and MSOD Strata: Treatment-Emergent Serious Adverse Events and Adverse Events of Special Interest Occurring in ≥5% of Patients in Any Group (Safety Population)*

| | Severe Patients | | | MSOD Patients | | |
|---|---|---|---|---|---|---|
| **System Organ Class**<br>**Preferred Term** | **Placebo Group (N=70)** | **Sarilumab 200 mg IV Group (N=140)** | **Sarilumab 400 mg IV Group (N=137)** | **Placebo Group (N=46)** | **Sarilumab 200 mg IV Group (N=95)** | **Sarilumab 400 mg IV Group (N=92)** |
| **SAEs** | | | | | | |
| SAEs — no. | 22 | 38 | 71 | 39 | 125 | 117 |
| Patients with ≥1 SAE — no. (%) | 11 (15.7) | 23 (16.4) | 30 (21.9) | 27 (58.7) | 67 (70.5) | 58 (63.0) |
| Infections and infestations — no. (%) | | | | | | |
| Covid-19 | 0 (0.0) | 2 (1.4) | 5 (3.6) | 5 (10.9) | 12 (12.6) | 9 (9.8) |
| Septic shock | 0 (0.0) | 1 (0.7) | 2 (1.5) | 0 (0.0) | 4 (4.2) | 5 (5.4) |
| Cardiac disorders — no. (%) | | | | | | |
| Cardiac arrest | 2 (2.9) | 4 (2.9) | 3 (2.2) | 6 (13.0) | 9 (9.5) | 9 (9.8) |
| Respiratory, thoracic, and mediastinal disorders — no. (%) | | | | | | |
| Respiratory failure | 2 (2.9) | 5 (3.6) | 4 (2.9) | 2 (4.3) | 10 (10.5) | 7 (7.6) |
| Acute respiratory failure | 1 (1.4) | 4 (2.9) | 4 (2.9) | 1 (2.2) | 3 (3.2) | 2 (2.2) |
| Vascular disorders — no. (%) | | | | | | |
| Hypotension | 0 (0.0) | 0 (0.0) | 1 (0.7) | 0 (0.0) | 2 (2.1) | 5 (5.4) |
| **AESIs** | | | | | | |
| AESIs — no. (%) | 8 | 24 | 45 | 54 | 81 | 90 |
| Patients with ≥1 AESI — no. (%) | 5 (7.1) | 20 (14.3) | 31 (22.6) | 28 (60.9) | 45 (47.4) | 51 (55.4) |
| Investigations — no. (%) | | | | | | |
| Alanine aminotransferase increased | 1 (1.4) | 5 (3.6) | 11 (8.0) | 9 (19.6) | 8 (8.4) | 14 (15.2) |
| Aspartate aminotransferase increased | 3 (4.3) | 6 (4.3) | 8 (5.8) | 12 (26.1) | 13 (13.7) | 14 (15.2) |
| Transaminases increased | 1 (1.4) | 3 (2.1) | 4 (2.9) | 4 (8.7) | 6 (6.3) | 3 (3.3) |
| Liver function test increased | 0 (0.0) | 2 (1.4) | 1 (0.7) | 3 (6.5) | 2 (2.1) | 5 (5.4) |
| Infections and infestations — no. (%) | | | | | | |
| Bacteremia | 0 (0.0) | 1 (0.7) | 2 (1.5) | 3 (6.5) | 4 (4.2) | 4 (4.3) |
| Pneumonia | 0 (0.0) | 0 (0.0) | 4 (2.9) | 0 | 5 (5.3) | 3 (3.3) |
| Staphylococcal infection | 0 (0.0) | 0 (0.0) | 0 (0.0) | 3 (6.5) | 6 (6.3) | 3 (3.3) |
| Pneumonia klebsiella | 0 (0.0) | 0 (0.0) | 0 (0.0) | 4 (8.7) | 3 (3.2) | 2 (2.2) |

*AESI denotes adverse event of special interest, IV intravenous, SAE serious adverse event.

#### Table S9. Association Between the *IL6R* Shedding Variant rs2228145:C and COVID-19 Outcomes*

| Phenotype | Study | Cases (no.) | Controls (no.) | Odds ratio (LCI, UCI) | P value |
|---|---|---|---|---|---|
| SARS-CoV-2–positive and hospitalized vs. population controls | COVID-19 Host Genetics Initiative Meta-Analysis | 12,888 | 1,295,966 | 0.96 (0.93, 0.99) | 0.007 |
| SARS-CoV-2–positive and hospitalized vs. not hospitalized | COVID-19 Host Genetics Initiative Meta-Analysis | 5,773 | 15,497 | 0.96 (0.90, 1.01) | 0.130 |
| SARS-CoV-2–positive and with severe disease vs. population controls | COVID-19 Host Genetics Initiative Meta-Analysis | 5,780 | 1,115,203 | 0.97 (0.92, 1.02) | 0.190 |

*LCI denotes lower confidence interval, UCI upper confidence interval.

We also considered two severity phenotypes, 1) hospitalization among PCR-confirmed SARS-CoV-2–infected individuals and 2) ventilation or death among SARS-CoV-2–infected individuals as compared with population controls, but these had less statistical power to detect associations. We found numerically lower odds of hospitalization among SARS-CoV-2–infected individuals (N=5773) compared with SARS-CoV-2–infected controls who were not hospitalized (N=15,497; TOR 0.96; 95% CI, 0.90 to 1.01; P=0.13) and lower odds of severe Covid-19 among SARS-CoV-2–infected individuals who required ventilation or died (N=5780) compared with population-level controls (OR 0.97; 95% CI, 0.92 to 1.02; P=0.19). These results were not statistically significant, suggesting larger sample sizes may be required. Altogether, these genetic findings are consistent with approximately 3% to 4% risk reduction for hospitalization and severe disease and provide support for IL-6 signaling blockade in Covid-19 management.
